# Early Parkinson’s Revealed by Unlocking Longitudinal Omics at Population Scale

**DOI:** 10.64898/2026.03.12.26348299

**Authors:** Chunmiao Feng, Idit Kosti, Yuelong Guo, Ying Wang, Nathan S. Watson-Haigh, Balint File, Nhi Hin, Tibor Nanasi, Jingyu Guo, Rad Suchecki, Rick Tearle, Kathleen Koborsi, Kim Dang, Rashi Saxena, Arnaud Teichert, Shalini Padmanabhan, Brit Mollenhauer, Samuel. M. Goldman, Tony Wyss-Coray, Karoly Nikolich, Scott Lohr, Benoit Lehallier

## Abstract

Many diseases begin developing years before symptoms appear^1–3^, yet biospecimens from these early stages are rarely available. We developed Chronos, a framework that uses privacy-preserving tokenization^4^ to link archived plasma samples with longitudinal clinical records, enabling the modeling of molecular trajectories across time. Starting with >100 million archived, routine-donation samples from 3 million plasma donors, we assembled a longitudinal Parkinson’s disease cohort and profiled 2,609 samples from 348 cases and 348 matched controls using four proteomics platforms, covering more than 25,000 proteoforms. We reproduced proteomic signatures from clinically-phenotyped cohorts and revealed early, coordinated alterations in a CXCL12, cell ratios to predict future diagnosis, achieving a maximum cross-validated area under the curve of 0.76 and replicated the findings in up to 5 independent cohorts. Chronos enables disease detection before clinical manifestation by prioritizing longitudinal molecular changes over symptoms, and provides a general framework to reconstruct chronic and acute disease trajectories from large plasma collections.

## Main text

The advancement of population-scale precision medicine hinges on the development of reliable, cost-effective, non-invasive biomarkers. While recent advances in multi-omics and AI-driven analytics have accelerated molecular discovery, translating these findings into clinically actionable tools remains a formidable challenge^5^. A primary bottleneck to further progress is the limited temporal depth of existing sample resources. Most available datasets fail to capture the biological complexity and molecular inflection points that occur years before clinical onset. Although prospective cohort studies are the current gold standard for mapping disease progression, they have notable limitations. They typically enroll participants only after symptom onset or in late prodromal stages and are often constrained by high attrition, substantial costs, and long study durations needed to capture relevant disease processes. Conversely, large-scale population studies such as the UK Biobank (UKB)^6^ provide breadth and scale but lack the individual-level temporal resolution needed to identify early predictive markers or to capture disease heterogeneity.

To overcome these barriers, we established Chronos, a scalable, repeatable framework for decoding the molecular mechanisms of disease across time (Fig. 1a). By integrating tokenized Real-World Data (RWD) with high-throughput molecular profiling in targeted and untargeted omics platforms^7,8^, Chronos transforms a repository of >100 million plasma samples into a molecular time machine for the study of health and disease (Supplementary Fig. 1a). The plasma repository from Grifols SA, built from more than 15 years of routine plasma collection at >300 donation centers in the US, serves as the biospecimen foundation for Chronos and provides longitudinal, population-scale representation across diverse demographic groups, including age, gender, race, and ethnicity (Supplementary Fig. 1b-h). The coverage of >50,000 ICD-10 codes enables the assembly of retrospective cohorts to investigate the molecular mechanisms underlying both common and rare diseases, including neurodegenerative, metabolic, cardiovascular disorders, and beyond (Fig. 1b).

**Fig. 1:**
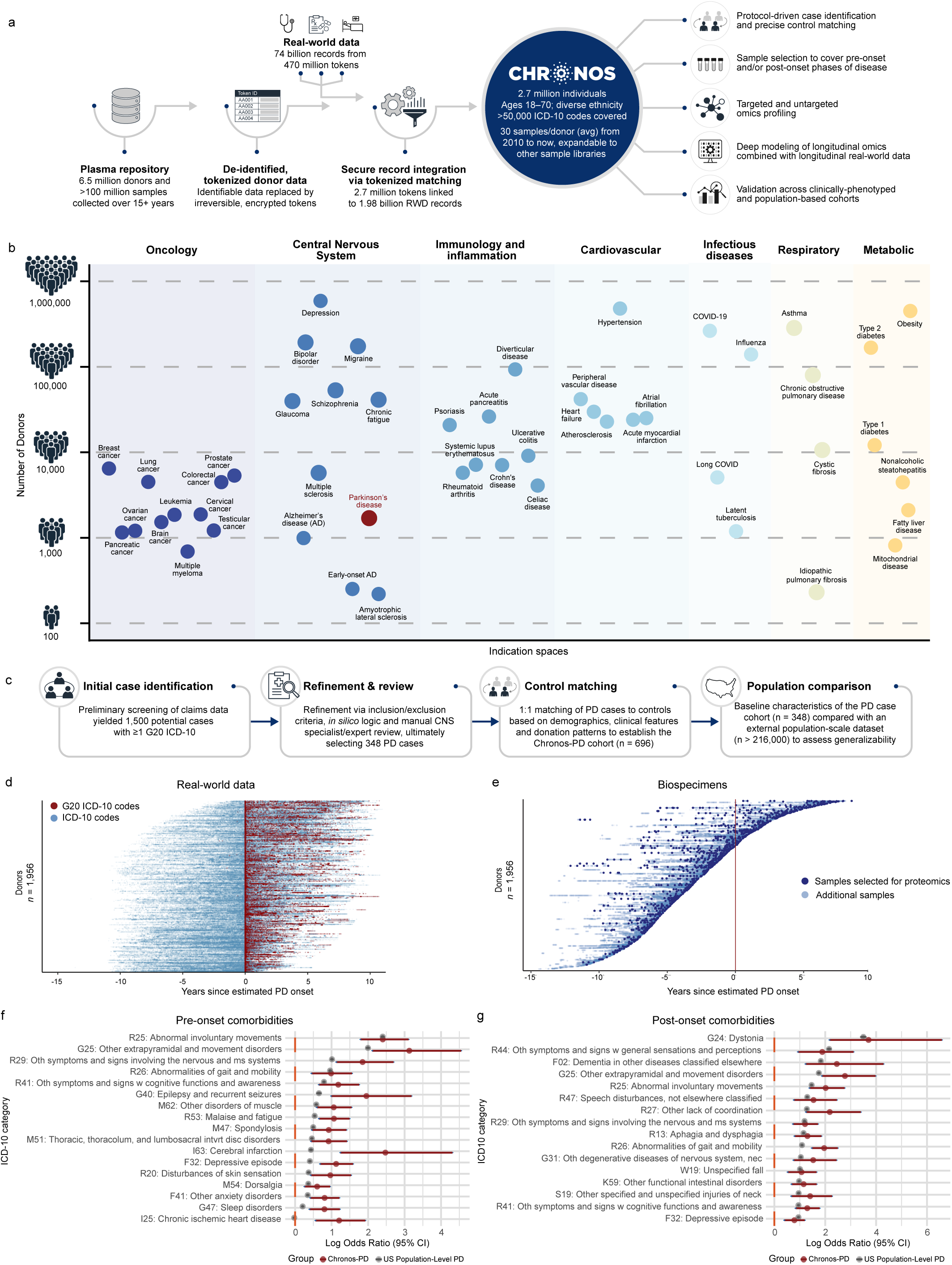
Identification and characterization of a representative Parkinson’s disease cohort from plasma donors. **a**, Overview of the Chronos workflow. The schematic illustrates the privacy-preserving tokenization of the donor registry, its secure linkage with de-identified real-world data (RWD), and downstream activities such as identification of study cohorts, case-control matching, and the retrieval of archived samples for omics profiling. **b**, Breakdown of the Chronos registry by disease category based on claims data. Distribution of donors with available plasma samples across seven major indication spaces, classified using linked RWD and ICD-10 coding. **c**, Flowchart reflecting the selection of PD cases and matched controls, including algorithmic screening and specialist review to ensure the integrity of the final cohort. **d-e**, Visualization of the PD case cohort’s longitudinal RWD records (dark red indicates PD-related claims) and samples selected for analysis (dark blue) relative to estimated PD onset. This temporal view highlights the density of data and samples available for the study period. **f-g**, Log odds ratios of leading comorbidities and symptoms, comparing the Chronos-PD case cohort pre- and post-onset and against a US population-level cohort (gray).

To demonstrate the potential of this framework, we focused on Parkinson’s disease (PD), the world’s fastest-growing neurodegenerative disorder^9^. Current PD diagnosis relies primarily on the assessment of cardinal motor symptoms (bradykinesia, tremor, and rigidity), which typically manifest only after substantial loss of dopaminergic neurons^1^. With no validated early-detection biomarkers or disease-modifying therapies available^10^, there is a critical need for objective, molecular biomarkers, particularly during the early preclinical stage when preservation of dopaminergic neurons may still be possible. In this study, we sought to elucidate the earliest molecular mechanisms of PD by “molecularizing” the most deeply sampled PD cohort to date, which currently includes >75,000 plasma samples from approximately 2,000 donors who developed PD, with >58,000 of those samples from the pre-diagnosis phase. From this larger PD resource, we selected a rigorously adjudicated subset with dense longitudinal sampling and matched controls to enable multi-platform proteomic profiling and trajectory modeling at high resolution.

### Defining a PD cohort from real-world data

To identify plasma donors who developed PD and enable precise control matching, we integrated the plasma donor registry of over 6.5 million individuals with RWD via secure tokenization and privacy-preserving record linkage (Methods). This integration linked 1.98 billion RWD records comprised of diagnostic codes, medication histories, and procedures with 2.7 million individuals with plasma samples usable for research (approximately 30 samples per donor on average). From this population, we applied a multistep cohort definition algorithm, yielding an initial selection of 1,500 putative PD cases (see Methods, Fig. 1c, and Supplementary Table 1). A further round of in-depth assessment by central nervous system (CNS) specialists was conducted to evaluate clinical plausibility and strength of supporting evidence (Supplementary Fig. 2a), yielding a final cohort of 348 individuals with the highest likelihood of PD. To obtain an objective estimate of the date of PD onset, a rule-based algorithm was applied integrating the temporal relationship between PD-related ICD-10 disease codes (G20) and relevant medication history (see Methods, Supplementary Fig. 2b). For the final cohort of 348 cases, an average of four longitudinal samples per individual were selected to provide the best coverage of PD evolution and maximize statistical power for characterizing molecular trajectories. Fig. 1d-e illustrates the extensive longitudinal RWD coverage and high-density sampling within the Chronos-PD cohort relative to estimated PD onset (557 encounters and 31 samples per individual on average) and highlights the 2,609 samples selected for molecular profiling in this study. Controls were selected from the same source donor collection by identifying individuals whose year of birth, race and ethnicity, gender, and comorbidity status (defined as the presence or absence of hypertension and type 2 diabetes) matched those of the cases. Matching was further refined based on sample collection timeframe and donation frequency (Supplementary Fig. 2c). For each case sample, a matching control sample was selected. This “Chronos Twin” strategy achieves near-perfect alignment of clinical and biospecimen features between cases and controls (Table 1), substantially minimizing confounding and thereby enhancing the precision and interpretability of molecular profiling analyses.

### Assessing Chronos-PD cohort representativeness

To confirm that the selected cohort exhibited expected PD symptoms and comorbidities, we calculated odds ratios (ORs) for three-digit ICD-10 codes in pre- and post-onset periods and compared those results with a similar analysis of a broader, representative population. In the pre-onset period, 17 comorbidities were significantly enriched in the PD case cohort after Benjamini Hochberg correction, including abnormal involuntary movements (OR = 11, 95% CI 6.0–22.4, q-value = 6.2 × 10⁻¹⁰), gait abnormalities (OR= 2.68, 95% CI 1.56-4.76, q-value = 0.033), sleep disorders (OR = 2.23, 95% CI 1.48-3.41, q-value = 0.015), and sensory disturbances (Fig. 1f, see full list in Supplementary Table 2). In the post-onset period, 64 comorbidities were significantly associated with our PD case cohort (Fig. 1g), with stronger signals for pre-onset symptoms such as gait and mobility abnormalities (OR = 7.09, 95% CI 4.36–12.09, q-value = 4.2 × 10⁻¹¹). Additional comorbidities associated with our PD case cohort included intestinal disorders (OR = 3.17, 95% CI 1.96–5.27, q-value = 3.5 × 10⁻⁴) and disorders of the urinary system (OR = 2.85, 95 % CI 1.79–4.65, q-value = 1.0 × 10⁻³) (see full list in Supplementary Table 2). Notably, we found that comorbidities were unevenly distributed across individuals (Supplementary Fig. 3a), with only five ICD-10 codes occurring in at least 25% of cases before estimated PD onset (R25 “abnormal involuntary movements”, M54 “dorsalgia”, R53 “malaise and fatigue”, F41 “other anxiety disorders”, and G89 “pain not classified elsewhere”). This uneven distribution persisted after onset, with eight ICD-10 codes reaching the same frequency threshold, underscoring the clinical heterogeneity of PD^11^ and the need for early, objective molecular biomarkers to predict risk, improve diagnosis, and guide therapeutic development. We then compared these findings with a similar analysis of de-identified RWD for a larger PD cohort of 216,336 individuals from the US population defined using stringent inclusion/exclusion criteria (see Methods). Sixteen of the 17 pre-onset comorbidities (94%) and all 64 post-onset comorbidities (100%) identified in the PD case cohort were also significant and directionally concordant in the US population PD cohort (Supplementary Fig. 3b). Together, these findings support the broader representativeness of our cohort and demonstrate that this methodology can be used to assemble cohorts that reflect the expected clinical features and comorbidity patterns of a complex, heterogeneous disease such as PD, while simultaneously providing biospecimen depth that enables unprecedented resolution for reconstructing disease trajectories.

### Proteomics profiling across four platforms

At present, no single proteomics platform comprehensively detects all proteins in plasma, nor the multitude of proteoforms they may adopt. Therefore, to maximize detection of PD-related molecular signals and to evaluate the performance and utility of various proteomic platforms, we profiled all 2,609 study samples using four complementary proteomics platforms (Fig. 2a-b, SomaScan v5, Olink Explore HT, Alamar Biosciences NULISAseq CNS Disease Panel 120, and Biognosys TrueDiscovery mass spectrometry referred to here as Somalogic-11k, Olink-HT-5k, AlamarBio-CNS-120, and discovery MS, respectively). While prior studies have shown the utility of these platforms individually for PD research (Supplementary Table 3), we performed a concurrent assessment of more than 25,000 assays, encompassing 14,000 unique proteins (Fig. 2b), producing the most comprehensive coverage of the PD plasma proteome to date.

**Fig. 2:**
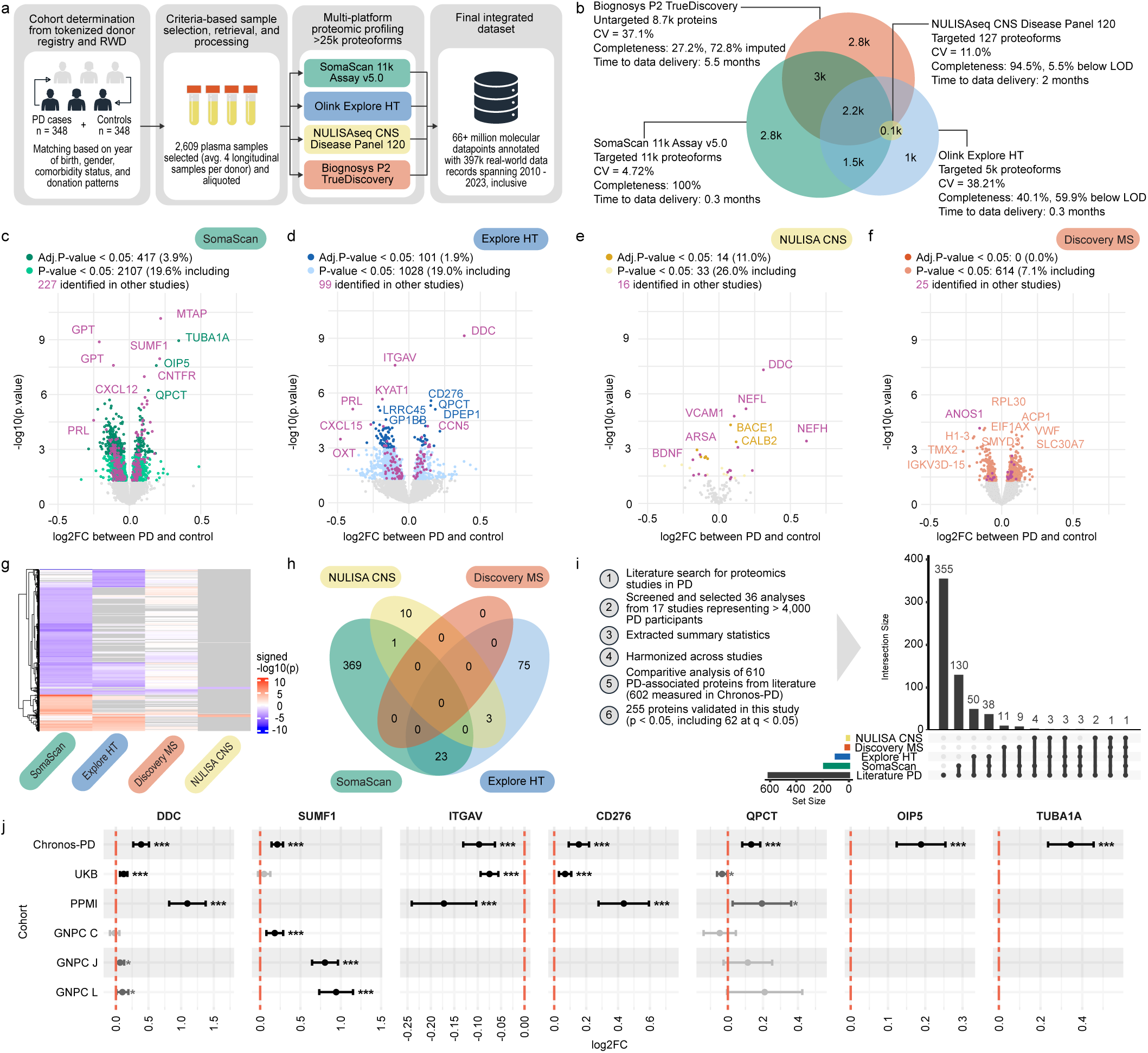
Comprehensive proteomic profiling of Parkinson’s disease. **a**, Flowchart of proteomics data generation from 2,609 plasma samples of 696 Chronos-PD donors, spanning four platforms. **b**, Venn diagram showing overlap of proteins measured across platforms and summary statistics for proteoform count, technical variation (CV), completeness, and data generation time. **c-f**, Volcano plots showing significant differences in protein levels (log2 fold change) between PD and controls for each platform, based on linear mixed models. New findings are shown in green for SomaScan (c), blue for Olink Explore HT (d), yellow for Alamar Biosciences NULISAseq CNS Disease Panel (e), and red for Biognosys P2 TrueDiscovery MS (f). Proteins in magenta signify findings consistent with previously reported PD studies. **g**, Heatmap of signed significance for PD-associated proteins across platforms. Gray indicates proteins not measured by a given platform, red-orange for increased proteins, and blue-purple for decreased proteins. **h**, Venn diagram of PD signals detected (q < 0.05) across platforms, showing overlap and platform-specific discoveries. **i,** Overview of literature search and comparison of Chronos-PD signals to published PD biomarkers, visualized by upset plot per platform. **j**, Forest plot of significant PD-associated proteins validated across Chronos-PD, UK Biobank, PPMI, and GNPC cohorts C, J, and L, showing estimated mean log2 fold change (log2FC) with 95% confidence interval (CI; error bars). Significance levels: *p < 0.05, **p < 0.01, ***p < 0.001.

Linear mixed model analysis on this dataset detected 417, 101, 14, and 0 proteoform signals showing significant changes in PD cases from the Somalogic-11k, Olink-HT-5k, AlamarBio-CNS-120, and discovery MS platforms, respectively (q-value < 0.05, Fig. 2c-2f, Supplementary Table 4). This represents 481 unique PD-associated proteoforms with overall concordance in direction and significance across platforms (Fig. 2g), although applying more stringent significance thresholds revealed noticeable differences in the number of PD-associated proteins identified per platform (Fig. 2h, Supplementary Table 5). Among the 481 PD-associated proteins, 23 were shared between Somalogic-11k and Olink-HT-5k (ADA2, CCL5, CCN5, CD300LG, CPXM1, CST5, CXCL3, ESM1, FBLN2, GMPR2, NAP1L4, OMD, PDGFRA, PRL, PTPN11, QPCT, ROR1, SCARF2, SERPINE1, SEZ6L, SLITRK1, SUSD5, TRIM25), 3 between Olink-HT-5k and AlamarBio-CNS-120 (DDC, GFAP, BACE1), and 1 common to Somalogic-11k and AlamarBio-CNS-120 (VCAM1). Somalogic-11k uniquely detected 369 (88% of 417) proteins, Olink-HT-5k 75 (74% of 101), and AlamarBio-CNS-120 10 (71% of 14), highlighting the complementarity of these platforms. Somalogic-11k showed the lowest technical variation (median CV = 4.7%), followed by AlamarBio-CNS-120 (11%), discovery MS (37.1%), and Olink-HT-5k (38.2%). Data completeness also varied substantially: >50% of the measurements from Olink-HT-5k and discovery MS were below the detection limit or missing, whereas AlamarBio-CNS-120 and Somalogic-11k were >90% complete. Somalogic-11k delivered four times more statistically significant PD signals than Olink-HT-5k (417 vs 101, q-value < 0.05). Although AlamarBio-CNS-120 produced fewer significant signals, consistent with its ultra-targeted design, it captured the highest proportion of PD-associated proteins relative to the number of proteins measured (11%, q-value < 0.05). Discovery MS detected no significant signals. These results highlight that, at multi-thousand-sample scale in plasma, discovery MS can be constrained by sensitivity and technical variability, supporting the use of single or combined affinity-based approaches for discovery, with MS best positioned for targeted validation and mechanistic follow-up.

### Expanding PD signatures from clinically-phenotyped cohorts

We next evaluated the consistency of our findings with known markers from clinically-phenotyped and population-based cohorts (Fig. 2i, Supplementary Table 3). First, we identified 610 PD-associated proteins reported across 17 prior studies (Supplementary Table 3). Of these, 602 (98.7%) were measured in Chronos-PD with 255 (41.8%) successfully validated with a p-value < 0.05 in our study and 62 with a q-value < 0.05. To substantiate these findings beyond published summary statistics, we obtained and reanalyzed raw data from established PD cohorts, including the Parkinson’s Progression Markers Initiative (PPMI) (project 9000), Global Neurodegeneration Proteomics Consortium (GNPC) cohorts C, F, J, L, and Q, and the population-based study UK Biobank (UKB). Chronos-PD showed strong concordance with five independent cohorts (PPMI, GNPC C, J, L and UKB; Supplementary Fig. 3a) in identifying proteins modulated by PD, though sign reversals were observed in GNPC cohort C across large protein sets, highlighting the need for systematic evaluation of normalization strategies and pre-analytical variability. Among the 481 unique PD protein signals identified in Chronos-PD (484 unique UniProt IDs), 307 (77%) of the 398 measured in at least one of these external cohorts were also found to significantly change in individuals with PD (p-value < 0.05). Many individual proteins displayed remarkably consistent patterns across studies (Fig. 2j), including well-established proteins associated with PD such as DOPA decarboxylase (DDC) (Supplementary Fig. 4b), and emerging ones such as integrin alpha-V (ITGAV)^12^ (Supplementary Fig. 4c), sulfatase-modifying factor 1 (SUMF1)^13^, and CD276 antigen (CD276). Further, proteins that were not measured in these external studies, such as tubulin alpha-1A (TUBA1A) and opa interacting protein 5 (OIP5), were first measured in PD in this study and emerge as new candidates, underscoring the importance of expanding proteomic coverage with advanced platforms (Fig. 2j).

Together, these findings demonstrate that the Chronos approach of integrating multiple proteomics platforms broadens molecular coverage, strengthens confidence in shared signals, and uncovers unique biology. This approach not only reproduced established proteomic signatures from clinically-phenotyped cohorts but also expanded them, providing a robust foundation for biomarker discovery during the preclinical phase of PD.

### Temporal patterns of proteomic change in PD

Our framework enables longitudinal characterization of molecular alterations and their temporal trajectories across all phases of PD. Many proteins showed undulating patterns throughout disease development (Fig. 3a), and restricting linear mixed modeling to pre-onset samples (n=1,748 pre-onset samples from 240 cases and 241 controls individuals) identified 14 proteoforms representing 12 proteins as significantly modulated before the estimated onset of PD (q-value < 0.05, Supplementary Table 4 and Fig. 3b). Proteome wide, this analysis focusing on pre-onset samples produced statistical significance patterns that were highly consistent with those obtained when using all samples (ρ = 0.59, p-value < 0.001; Fig. 3b), supporting the robustness of the early signals identified. Conversely, five proteins including SUMF1, OIP5, TUBA1A, GPT, and DDC showed minimal change before onset and marked shifts during the peri- and post-onset periods, suggesting involvement in later disease phases or reflecting downstream consequences of treatment.

**Fig. 3:**
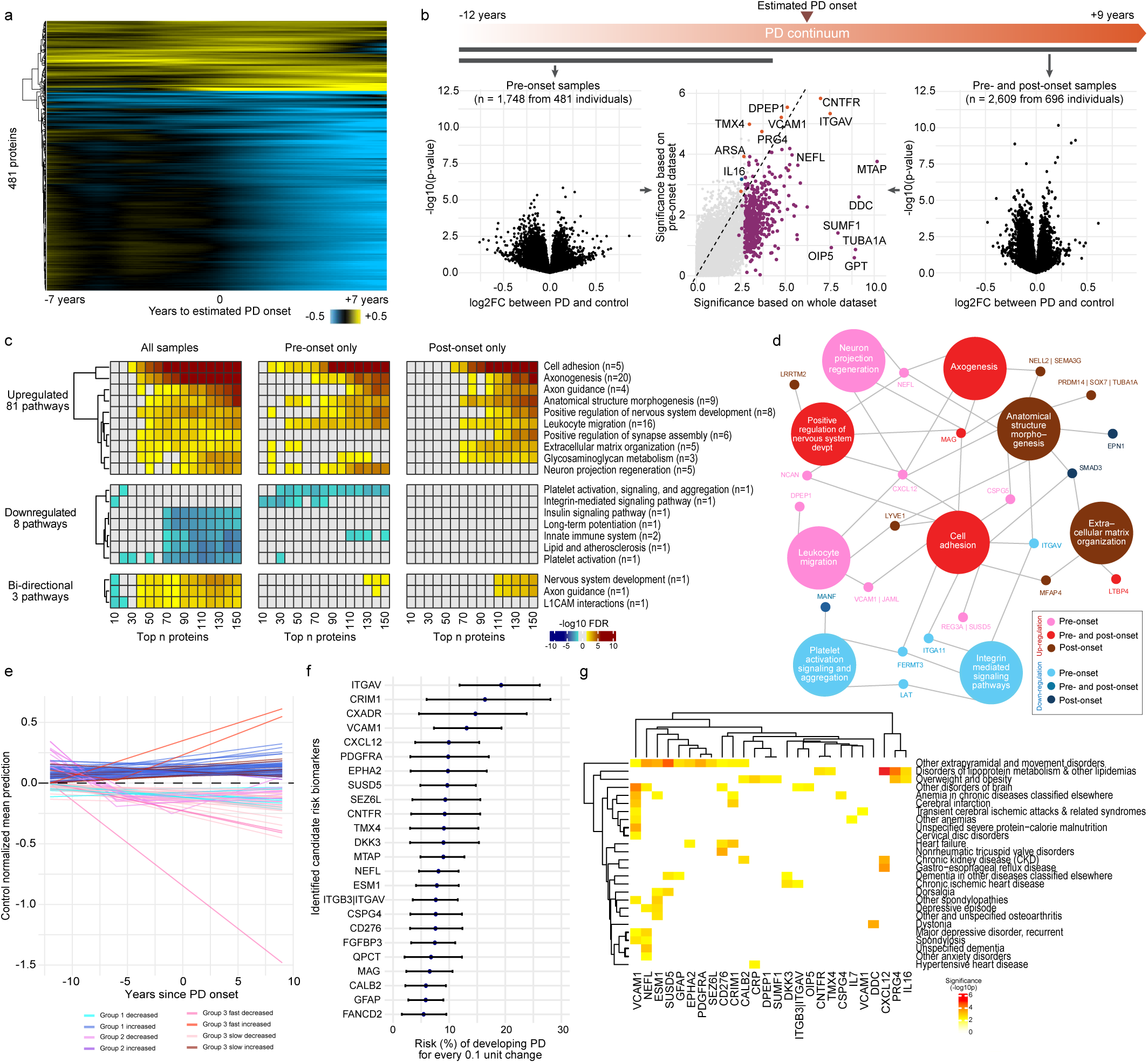
Longitudinal molecular insights into the earliest signals of Parkinson’s disease. **a**, Heatmap of PD protein trajectories estimated by locally estimated scatterplot smoothing (LOESS), standardized to control levels. Blue indicates decreased, yellow increased, and black unchanged protein levels. **b**, Volcano plots comparing PD signals from pre-onset samples (n = 1,748) and all samples (n = 2,609), with color coding for proteins significant in both (red), only pre-onset (blue), or only whole sample analyses (purple). **c**, Key enriched pathways among PD signals from pre-onset, post-onset, and all samples, detected by SEPA. Row blocks indicate upregulated, downregulated, or bi-directional pathway sets; cell colors represent signed -log10(FDR), brown for upregulated pathways, blue for downregulated pathways. Redundant pathways are collapsed into a single line. **d**, Network visualization of top pathway-protein associations. Rectangles denote pathways, circles denote proteins, with edges indicating membership. Node color encodes pre-onset and post-onset regulation. **e**, Protein trajectories identified based on linear, non-linear mixed models, joint modeling of protein levels, and time to PD onset. Trajectories are visualized as control-normalized estimates from 12 years before to 9 years after PD onset. **f**, Top candidate PD risk biomarkers from joint Cox regression and linear mixed models, showing estimated mean risk and 95% confidence interval (CI) for proteins with >5% risk per 0.1 unit change (p < 0.005). **g**, Heatmap of significant associations between PD-associated comorbidities and early/risk biomarkers (filtered to p < 0.01), with color representing statistical significance.

To assess whether medication contributed to these changes, we conducted a focused analysis of levodopa effects by comparing samples collected after a levodopa prescription claim to those without a nearby levodopa prescription claim. This analysis replicated previously reported levodopa-associated increases in plasma DDC levels^14,15^ and suggests that TUBA1A, OIP5, and GPT may also be influenced by levodopa treatment (uncorrected p < 0.05, Supplementary Fig.4d). Although power was limited (n = 54 cases for this focused levodopa analysis), the large effect sizes observed for other proteins (|Cohen’s d| ≥ 0.8 for 261 proteoforms) point to a substantial impact of levodopa on the plasma proteome. This observation highlights the need for future studies to account for these dynamics with the aim of potentially disentangling the effects of levodopa and other PD medications.

Among the strongest pre-onset signals (Supplementary Table 4) were alterations in ciliary neurotrophic factor receptor (CNTFR), dipeptidase 1 (DPEP1), integrin subunit alpha V (ITGAV), vascular cell adhesion molecule 1 (VCAM1), thioredoxin-related transmembrane protein 4 (TMX4), proteoglycan 4 or lubricin (PRG4), and Arylsulfatase A (ARSA) (q-value < 0.05). ARSA acts as a genetic modifier of PD by functioning as a molecular chaperone that directly binds α-synuclein in the cytosol, thereby inhibiting its toxic aggregation and cell-to-cell propagation^16,17^. CNTFR, a neurotrophic receptor supporting dopaminergic neuron survival, may represent a compensatory mechanism during the prodromal stage. DPEP1 promotes ferroptosis, a form of regulated cell death, through glutathione depletion and lipid peroxidation, thereby exposing oxidative vulnerability which is a well-established hallmark of PD^18,19^, and suggesting that this vulnerability begins to change early in the disease course. VCAM1 and ITGAV molecules regulate immune cell trafficking across the blood–brain barrier and link vascular inflammation to synaptic dysfunction, a mechanism increasingly recognized as pivotal in PD pathogenesis. Plasma VCAM1 levels correlate with PD severity^20^, and independent cohorts confirm that VCAM1 and ITGAV proteins are altered in plasma prior to PD diagnosis^12,21^. Building on these protein-level findings, pathway analysis revealed stronger enrichment among pre-onset proteins, compared to those observed post onset, characterized by upregulation of cell adhesion molecules, neuronal and axonal pathways, and leukocyte activation, alongside downregulation of integrin signaling, innate immune responses, and platelet function (Fig. 3c-d, expanded results in Supplementary Fig. 5a and Supplementary Table 6). Although individual protein level changes were less pronounced before PD onset, these proteins mapped to more specific and biologically coherent pathways. This suggests that early mechanistic shifts precede clinical onset and signal a transition from localized perturbations to a progressively more widespread and heterogeneous state as the disease advances and downstream biological systems become engaged. This pattern aligns with the recognized clinical heterogeneity of PD.

### Biomarker trajectories and risk signatures

To deepen our understanding of these findings and explore potential applications of the biomarkers we identified, we applied non-linear mixed models (linear spline and natural cubic spline, see Methods) to detect proteins exhibiting non-linear trajectories and selected the best-fitting model for each protein based on the Bayesian Information Criterion (BIC). Integrating results from linear mixed models, non-linear mixed models, but also joint modeling coupling longitudinal protein trajectories with time relative to onset, we identified three distinct biomarker groups based on patterns of change (Fig. 3e, Supplementary Fig. 5b, Supplementary Table 7): one group which shows early linear change (group 1, 10y+ before onset, n=47 proteoforms), a second group which shows inflection years before onset (group 2, n=12 proteoforms), and a third group which exhibits late linear and non-linear change (mostly pre- and post-onset, group 3, n=36 proteoforms). These groups suggest distinct clinical utilities. Early linear profiles may serve as predictive or risk markers identifying individuals at heightened susceptibility, while inflection years before onset may be useful for early diagnosis and stratification, and late linear and nonlinear profiles may be more relevant for monitoring disease progression or assessing treatment response. Although these roles are not mutually exclusive, mechanistic context and biological validation will ultimately guide their clinical deployment.

Because this study offers a unique opportunity to examine early molecular indicators of disease development, a central objective was to determine whether risk biomarkers with predictive utility for PD onset could be identified. Based on joint modeling of molecular profiles and survival data, we identified dozens of candidate biomarkers (Fig. 3f, Supplementary Table 8). ITGAV showed the largest effect, increasing PD risk by up to 20% per 0.1-unit rise, while VCAM1, CXCL12, CRIM1, and CXADR were associated with 10–15% changes in risk. CXCL12/CXCR4 signaling activates FAK/Src/Rac1 pathways involved in microglial migration and α-synuclein-induced neuroinflammation^22^, whereas VCAM1 and integrins facilitate leukocyte adhesion and blood-brain barrier integrity^23^. Together, these processes may promote a pro-inflammatory environment that accelerate α-synuclein aggregation and dopaminergic neuronal loss. Dysregulation of this axis could amplify leukocyte trafficking and inflammatory cascades, supporting a model in which neurovascular immune interactions shift years before clinical onset.

To assess the robustness of these candidate risk biomarkers, we tested their associations with PD-associated chronic comorbidities (Fig. 3g, Supplementary Table 9) and other conditions (Supplementary Fig. 6, Supplementary Table 10). These results indicate that overall, the candidate risk biomarkers are not strongly driven by comorbidities. Instead, the comorbidities that do show associations reinforce biological plausibility (Fig. 3g). For example, the concentration of VCAM1 signal within neurological and cerebrovascular comorbidities supports its role at the neurovascular immune interface. In parallel, metabolic conditions such as hyperlipidemia, obesity, and weight disorders were each associated with multiple early biomarker candidates (Fig. 3g), reinforcing the link between metabolic dysregulation and PD pathophysiology^24^. The strong overlap with extrapyramidal and movement disorders further validates these signals within the shared clinical spectrum. Collectively, these findings provide strong evidence that these candidate biomarkers reflect early pathobiological activity in PD development and highlight mechanistic pathways relevant to PD risk. Our results converge on a preclinical axis linking CXCL12, VCAM1, and integrins, outlining early neuroimmune crosstalk and highlighting testable targets along a path toward intercepting PD before clinical onset.

### Early prediction of PD onset

To assess whether early molecular changes carry predictive value based on a single sample per individual, we first tested the ability of single proteins to classify future PD cases using logistic regression on samples collected 1 to 3 years before estimated onset (164 paired cases and controls). However, this approach yielded only modest discrimination, with maximum area under the curve (AUC) < 0.70 for single proteins (CV AUC = 0.68 for CD3E). Machine learning models (XGBoost and elastic net) that combined multiple proteins performed poorly, with AUCs near 0.6 when affinity proteomics platforms were evaluated separately and falling to 0.52 when combined, likely reflecting the challenges posed by limited sample size and high feature dimensionality for ML algorithms.

To address these limitations, we investigated protein ratios^25^. Ratios compress high-dimensional discovery into assayable paired readouts that may be more portable across platforms and pre-analytical variability^26^. Pairwise protein ratios were systematically generated from affinity platform assays, resulting in over 131 million within- and between-platform pairs (Fig. 4a). We identified ratios with clear predictive utility (AUC > 0.7, max AUC of 0.76) and prioritized 193 ratios based on consistent discrimination, requiring an AUC > 0.6 in at least three pre-onset two-year sliding windows. These ratios increased statistical significance and reduced confidence intervals compared to their individual protein components (Supplementary Fig. 7).

**Fig. 4:**
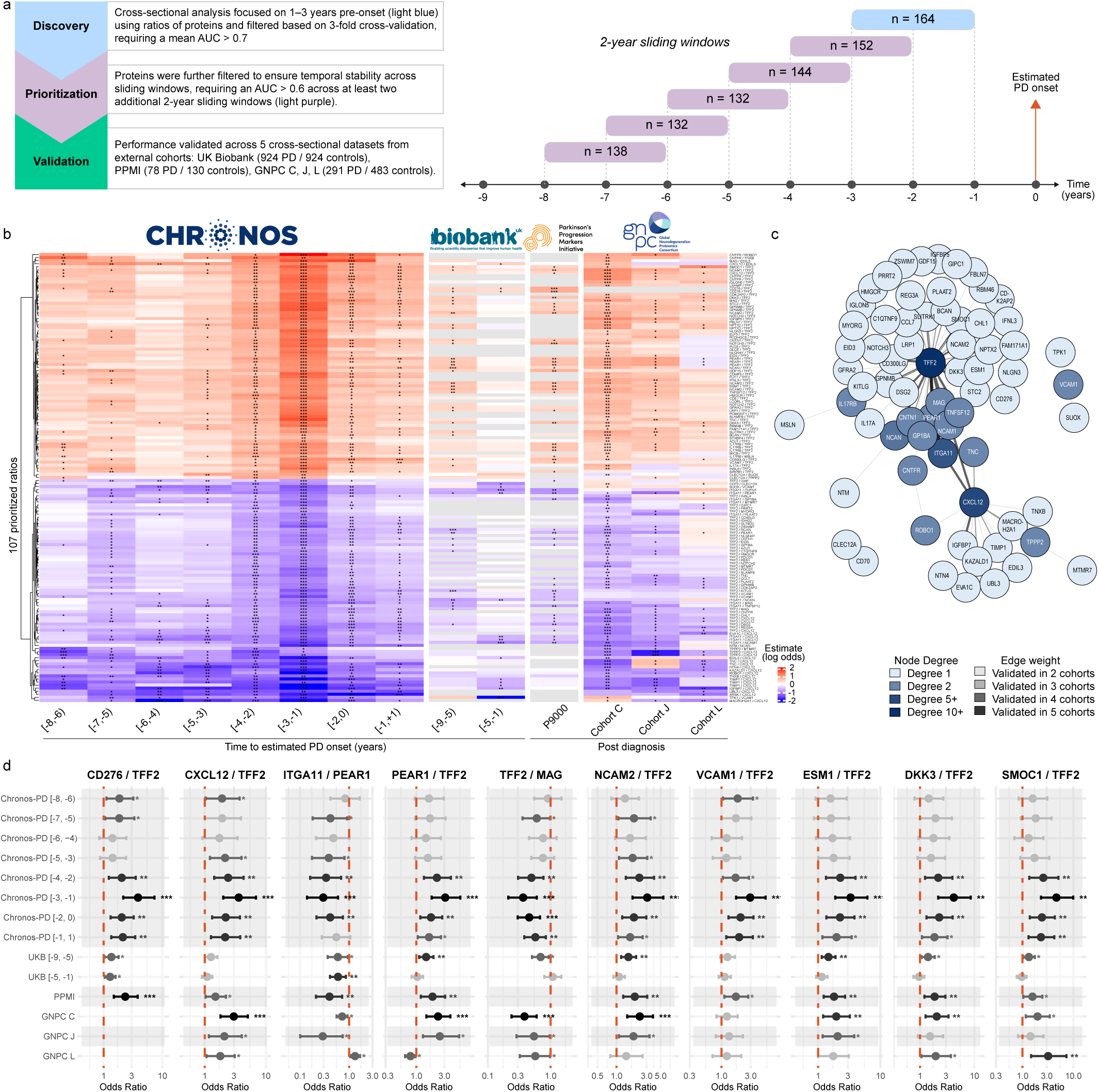
Identification and validation of protein ratios predictive of prevalent and incident Parkinson’s disease. **a**, Overview of discovery and validation strategy for protein ratios. Logistic models were fitted in 2-year sliding windows, with performance assessed by 3-fold cross-validation AUC (light blue) in the [-3,-1) window. Ratios with AUC > 0.7 were further evaluated in neighboring windows (light purple), and five external cohorts. **b**, Heatmap of log-odds estimates for 107 prioritized protein ratios significant in at least one external cohort. Columns represent 2-year windows in Chronos-PD (aligned to estimated PD onset), 4-year windows in UK Biobank, and post-diagnosis cohorts (PPMI, GNPC C, J, L). Rows are ratios; colors indicate effect estimates (log-odds of PD per one-unit increase in ratio), with asterisks for significance levels (***p < 0.001, **p < 0.01, *p < 0.05). **c**, Network plot of proteins consituting the prioritized ratios validated across at least two external cohorts, with nodes for proteins and edges for ratio pairs. Node color reflects connection degree (number of connections a node has); edge color reflects validation strength (times validated in external cohorts). **d**, Forest plot of odds ratios (ORs) for selected protein ratios across cohorts. points and error bars denote ORs and 95% CIs; asterisks denote statistical significance (***p < 0.001, **p < 0.01, *p < 0.05).

We then assessed these ratios in external cohorts to substantiate our findings. Of the 193 ratios prioritized in the Chronos-PD study, 39 (20.6%) could not be assessed in any external cohort because the component proteins were not measured in these datasets. The remaining 154 were tested in at least one of the PPMI, UKB, or GNPC C, J, and L cohorts with 108 (70%) validated in at least one external cohort (p < 0.05, Fig. 4b). Furthermore, a substantial subset of ratios demonstrated robust reproducibility, with 18 ratios validated in ≥ 4 external cohorts. Among these, the ratio of Platelet endothelial aggregation receptor 1 (PEAR1) to Trefoil factor 2 (TFF2) and the ratio of Integrin alpha-11 (ITGA11) to TFF2 emerged as particularly robust candidates, achieving statistical significance across 5 independent cohorts. Notably, the external cohorts were profiled using different affinity proteomic platforms (SomaLogic and Olink), suggesting that protein ratios can mitigate both analytical and pre-analytical variation and improve reproducibility across heterogeneous datasets. Taken as a whole, the external validation across at least two external cohorts identified a robust network of 75 (39%) protein ratios with TFF2, ITGA11, CXCL12, and NCAN emerging as central nodes involved in more than three ratios each (Fig. 4c). Although TFF2 lacked predictive value as an individual marker in linear mixed models, its predictive utility emerged when incorporated into composite ratios. Biologically, TFF2 is secreted by gastric and intestinal mucosa and is closely linked to gut epithelial repair^27^, with the growing evidence for gut involvement in PD pathogenesis^28^ supporting its relevance. In parallel, we identified CXCL12 as a PD risk biomarker (Fig. 3b), with the CXCL12/TFF2 ratio validated across four independent cohorts. Mechanistically, this association may reflect TFF2 acting as partial agonist of CXCR4^29^, thereby modulating CXCL12 signaling and downstream neuroimmune pathways. The recurrent involvement of TFF2 across multiple validated ratios, together with its functional interplay with CXCL12, points to a coherent mechanistic signal capturing coordinated regulation across immune, gut brain, and adhesion related processes.

Altogether, this study provides key insight into molecular changes preceding clinical PD and demonstrates that specific protein ratios markedly enhance early disease prediction, with consistent validation across independent cohorts. By reflecting biologically coherent changes rather than isolated signals, protein ratios provide a reproducible, mechanistically grounded approach for risk stratification in the preclinical phase. The Chronos framework establishes a scalable approach for biomarker-driven early detection and paves the way for future clinical translation in PD and other complex disorders.

## Discussion

Identifying molecular inflection points in the transition from health to disease requires both massive scale and a deep retrospective window, which traditional prospective cohorts rarely achieve. By combining routine plasma collection at population scale (>100 million samples from a donor registry of 6.5M individuals) with privacy-preserving linkage to real-world clinical data, the Chronos framework enables molecular profiling up to 15 years before and 10 years after diagnosis. This scale and temporal depth provide a unique opportunity to discover early biomarkers and therapeutic targets. In contrast to large biobanks such as UK Biobank^30^ or research cohorts such as the Parkinson Progression Markers Initiative^31^, Chronos offers dense longitudinal biospecimen coverage across both pre- and post-onset phases (Supplementary Fig. 8a).

In PD, we recapitulate molecular changes observed in clinically-phenotyped cohorts (Fig. 2c-f,i, Supplementary Fig. 4a-b,d), confirming the generalizability of findings using this framework, and expand them to the preclinical phase of the disease. We highlight pre-diagnosis molecular changes in the CXCL12 - cell adhesion molecules - integrin axis (Fig. 3), which regulates neuroimmune interactions and blood-brain barrier integrity providing actionable targets for early disease interception. Several proteins from this axis were reported individually in prior studies; here, we demonstrate that they form a coherent functional network that begins to change years before the clinical manifestation of PD. Because Chronos spans all phases of PD, from the preclinical stage to post-diagnosis, we were able to detect candidate biomarkers with distinct temporal trajectories and potential clinical utility (Fig. 3e, Supplementary Fig. 5b).

Identifying biomarker candidates appearing prior to clinical onset is of critical importance because they may provide a window for early intervention and risk stratification before irreversible neurodegeneration occurs (Fig. 3f, Fig. 4). We uncovered and prioritized 193 protein ratios with strong potential prognostic value (CV AUC max of 0.76) and validated them across up to five independent cohorts profiled using two distinct affinity-based proteomics platforms. These AUC values are notable given the prevalence of the misdiagnosis of PD^32^ and, importantly, when viewed in the context of the Alzheimer’s disease diagnostic development trajectory, where early plasma studies of total tau showed similarly modest AUC performance and meaningful clinical utility emerged only after assays targeting specific p-tau proteoforms were introduced. The ratios identified here already demonstrate encouraging prognostic performance at the discovery stage, with replication across platforms and cohorts underscoring the robustness of these signals and the compelling potential of select ratios to serve as early PD biomarkers. At the same time, the large number of strong candidates highlights the need for systematic prioritization and continued assay development to enable clinical translation.

Plasma proteomics provides access to the functional layer linking genetic variation to clinical phenotypes, and this rapidly advancing field now illuminates biological mechanisms that were previously inaccessible. Our findings highlight the complementarity of affinity proteomics platforms in reconstructing complex biological systems and better resolve the multifactorial nature of PD. The complex interplay between CXCL12 - cell adhesion molecules and integrins in early PD would not have been captured by a single proteomics platform. Indeed, CXCL12 was identified exclusively with Somalogic-11k, as it was not measured by other affinity platforms (Supplementary Fig. 8b); VCAM1 was detected in both Somalogic-11k and Alamar, but low signal in hundreds of Olink-HT-5k samples precluded PD signal detection (Supplementary Fig. 8c); and distinct integrins were identified across Somalogic-11k and Olink-HT-5k. A substantial fraction of PD-associated proteins identified here (15% for Olink-HT-5k and 21% for SomaLogic-11k) were absent from earlier versions of these platforms, underscoring how rapidly affinity technologies expand their coverage and the value of reprofiling samples with state-of-the-art assays to capture newly measurable biomarkers. In contrast, discovery MS reaches practical limits at biomarker-discovery scales above one thousand samples and may be better suited for targeted validation of affinity-based signals and for in-depth mechanistic follow-up. Importantly, multiple longitudinal samples markedly increased statistical power, yielding up to a 15-fold increase in significant associations compared with a cross-sectional design with the same number of participants. Significant associations plateaued with three to four samples and decreased slightly with five, suggesting that deep longitudinal sampling could offer a practical balance between statistical power and false positives, though further validation is needed (Supplementary Fig. 8d). The Chronos framework opens the door to scalable research across diverse understudied populations (27% of the individuals in the plasma donor population used in this study are African American, 20% Hispanic/Latino, and individuals aged 18 to 70, Supplementary Fig. 1), helping to close global research gaps created by decades of Eurocentric data^33^. Because clinical characterization and disease progression often differ between research cohorts and real-world populations^34^, the ability to profile molecular changes directly within communities provides a powerful and complementary perspective to prospective studies. Although the retrospective design and reliance on RWD for disease ascertainment and onset estimation introduce diagnostic uncertainty, rigorous multi-step cohort definition and expert review help mitigate these limitations. Beyond addressing these considerations, we show that Chronos functions as a bridge between real world populations and traditional research cohorts by demonstrating that molecular signatures discovered in the real-world population validate independently in multiple clinically-phenotyped cohorts, allowing their clinical relevance to be directly evaluated (Fig. 2c–f,i; Supplementary Fig. 4a–c). The availability of detailed health records through RWD supports comprehensive tracking of comorbidities, medication exposure, and clinical milestones over time, facilitating systematic dissection of molecular signals (Supplementary Fig. 4d and 6). This approach can also be used to embrace the diagnostic heterogeneity inherent in PD, where misclassification and co-pathologies are common, and supports the molecular characterization of PD and overlapping neurodegenerative diseases beyond ICD-10 labels toward biologically grounded disease signatures. Indeed, the >75,000 available samples from 2,000 individuals with PD can be immediately compared with thousands of individuals with frontotemporal dementia, dementia with Lewy bodies, Alzheimer’s disease, multiple system atrophy, and secondary Parkinsonism (Supplementary Fig. 9).

Together, our results demonstrate that large-scale integration of real-world and population-based biospecimens and molecular profiling with longitudinal RWD provides a multidimensional view of disease initiation. This framework extends to virtually any disease (Fig. 1b). It is well suited to conditions with long preclinical phases, including diabetes and cancer, and the high frequency of plasma donation in the United States, up to two times per week, also enables investigation of acute disease mechanisms such as cardiovascular or autoimmune disorders. We strongly encourage adoption of this framework beyond the current sample library, including national plasma collection efforts, to accelerate early disease detection and precision medicine at scale.

## Methods

### Study Design and Ethical Approval

This retrospective, longitudinal, case-control study was approved by Advarra’s Institutional Review Board (IRB Protocol #Pro00081762). The requirement for informed consent was waived on the basis of minimal risk to donors and the use of de-identified data and archived biospecimens.

### Data Sources and Record Linkage

#### Sample Repository

Samples utilized in this study originated from routine source plasma collection conducted by Grifols’ network of over 300 licensed plasma donation centers across the United States (US). Grifols, a global healthcare company and manufacturer of plasma-derived medicines, operates these centers under standardized procedures that adhere to all applicable regulations (e.g., 21 CFR 640.60 and 21 CFR 630.15), ensuring donor consent, health and safety, and plasma quality. Routine plasma collection is performed via plasmapheresis^35–37^ using sodium citrate as an anticoagulant. Donors are permitted up to two donations per week, with a minimum of two days between sessions. From each donation, three samples are routinely extracted: two for mandatory testing (Nucleic Acid Testing, NAT, and serology) and one backup sample (2.5 mL or 3 mL plastic blood collection tube with no additives). The backup samples are stored frozen (at or below -20°C) within 30 minutes of collection, and when no longer needed for their primary use (e.g., re-testing, etc.), may be maintained in long-term storage at -30°C for potential research, development, or other approved use. This sample repository comprises over 100 million samples contributed by 3 million unique donors from Grifols’ registry of 6.5 million and is expanding by >10 million additional samples per year. Potential effects of storage duration or repeated donation on proteomic profiles were addressed through Chronos Twin matching, which aligns donation timing and donation frequency between cases and controls.

### Real-World Data Integration and Linkage

To establish a comprehensive longitudinal health record for the plasma donors, real-world data (RWD) was integrated with the plasma sample information. Real-world data were sourced from Kythera Labs Open Claims. Data integration was achieved using privacy-preserving record linkage facilitated by tokenization via Datavant’s software. Personally identifiable information (PII) for the plasma donors was accessed by authorized study personnel to generate donor-specific, encrypted tokens. Datavant’s platform then securely matched corresponding open claims data from Kythera using the combination of Datavant tokens 1 and 2. The linkage process successfully matched 5.25 million of the 6.5 million plasma donors from the Grifols registry to the Kythera Labs data, resulting in a linkage rate of 81% for the primary claims database. That population was later linked to plasma samples that can be used for research, resulting in 2.7M donors. A similar approach was used to incorporate open claims and electronic health records (EHR) from Clarivate and PurpleLab into the expanded RWD Chronos resource shown in Fig. 1b,d-e; however, these two additional sources were not available when the PD cohort for this study was assembled.

### Study Population

#### Identification of Parkinson’s Disease Cohort

We identified 348 PD cases using a multistep process combining automated algorithmic screening and manual expert adjudication. Following the removal of donor PII and integration with de-identified, tokenized RWD, a minimum set of filtering criteria was applied to identify individuals who met the following requirements: (1) availability of at least one archived plasma sample for research; (2) evidence of at least three distinct clinical encounters with valid diagnosis dates; and (3) at least one recorded PD diagnosis code (specifically, ICD-9 code 332.0 and/or ICD-10 code G20, including all associated child codes) (see Supplementary Table 12 for full list). The final PD cohort was assembled from five sub-cohort definitions (shown in Supplementary Table 1) with inclusion and exclusion rules based on diagnoses codes (Supplementary Table 12), medications (Supplementary Table 13), and PD symptoms (Supplementary Table 14). The sub-cohorts are described by order of placement in PD cohort.

The first sub-cohort, subsequently referred to as *Established sub-cohort*, included 125 individuals and was defined based on the presence of two or more PD diagnosis codes; two PD medications prescriptions that were sixty days or more apart; absence of any prescription claims for antipsychotic medications (exclusion medications, Supplementary Table 13); and absence of exclusion codes representing other CNS conditions with phenotype that can be similar to PD (Supplementary Table 12). This sub-cohort applies strict criteria and excludes confounding conditions or medications, improving diagnostic specificity.

The second sub-cohort, subsequently referred to as *Symptomatic sub-cohort*, included 105 individuals and was defined using five or more PD diagnosis codes across six or more months, together with five PD symptom codes (Supplementary Table 14) across six or more months. This sub-cohort captures individuals with persistent PD-related symptoms over time.

The third sub-cohort, subsequently referred to as *Dx/Rx sub-cohort*, included in 77 individuals and was defined using one or more PD diagnosis codes, and two or more PD medications prescriptions that were sixty days or more apart. This sub-cohort provides a broad set of PD cases identified through diagnosis and medication patterns, ensuring inclusion of individuals with clinical and pharmacological evidence of PD.

The fourth sub-cohort, referred to as the *DBS (deep brain stimulation) sub-cohort*, included five individuals and was defined using DBS procedural codes from the Medtronic manual together with PD diagnosis codes. DBS can be performed for multiple indications, including conditions unrelated to PD, such as epilepsy (ICD 10 code G40 and related child codes) and obsessive compulsive disorder (ICD 10 code F42 and related child codes)^38^. Other conditions, including dystonia (G24) and other extrapyramidal and movement disorders (G25), may represent manifestations of PD. To ensure that DBS was performed in the context of PD, we selected only individuals with a G20 diagnosis code recorded on the same day as the DBS procedure. This criterion allowed us to distinguish DBS procedures performed for PD from those performed for other indications. This sub-cohort represents advanced PD cases requiring surgical intervention. The fifth sub-cohort, subsequently referred to as *classifier sub-cohort*, included 36 individuals and was defined using XGBoost classifiers (implemented with the *xgb.train()* function from the *xgboost* v1.7.8.1 R package) applied to RWD from plasma donor data without any sample restrictions. Two classifiers were trained: one to distinguish *established PD* cases (n = 634, defined as described above) from controls with no G20 diagnosis and no PD medications (n = 634 randomly sampled from 4,626,340 individuals, exclusion medications listed in Supplementary Table 13), and a second to distinguish the same established PD cases (n = 634) from a non-PD cohort (n = 634 randomly sampled from 123,662 individuals). The non-PD cohort was defined as donors with zero PD inclusion claims (Supplementary Table 12), zero PD inclusion medications (Supplementary Table 13), and at least 10 unique PD exclusion medications (Supplementary Table 13) claims spanning a minimum of one year. These exclusion medications are associated with symptoms that can resemble PD, although these donors were never diagnosed with PD. Individuals from the majority classes were selected by random sampling, and the sampling procedure followed by xgb model building were repeated three times to improve the confidence of the predictions. This *classifier sub-cohort* consisted of donors with a predicted probability greater than 0.9 of being a PD patient according to both classifiers. This sub-cohort leverages machine learning to identify high-probability PD cases from real-world data, expanding the cohort with predictive precision beyond rule-based criteria.

Except for the *established subcohort*, which met stringent algorithmic criteria and was not reviewed manually, all candidate cases underwent independent review using an internal revision system, which displays all available RWD and donation information (Patient Event Visualization, Supplementary Fig. 2a), by at least two CNS specialists. Instances of disagreement were escalated to a PD expert for an additional level of review and adjudication.

### Estimated Date of PD Onset

The estimated PD onset date was assigned using ICD-9/10 diagnosis codes (332.0, G20 and child codes) and PD medication records (PDm, Supplementary Table 13). The full decision flow is shown in Supplementary Fig. 2b. Among the 348 PD cases, 130 (37%) had no PDm records (or no medications records); for these donors, the first G20 code was assigned as the onset date. For the remaining 218 donors (63%) with PDm records, onset was defined based on the interval between the first G20 code and the first PDm. For 154 donors (71%) where the first PDm occurred no more than one year before the first G20 code, onset was set to the earliest of these two dates. For 64 donors (29%) where the first PDm occurred more than one year prior to the first G20 code, onset was determined through expert review, considering clinical context and medication stability.

In these cases, onset was assigned to either the first G20 code, the first PDm date, or the first date suggested by the PD expert. This approach yielded a more accurate onset estimate than relying solely on the first ICD-9/10 diagnosis by integrating diagnostic and treatment timelines and applying expert adjudication for ambiguous cases.

### Control Matching Process for Chronos Twins

We performed a 1:1 case-control matching to reduce demographics, clinical and pre-analytical confounders. To this end, potential controls were selected among individuals meeting all study eligibility criteria described above, resulting in a pool of 1.66 million after excluding those with PD diagnosis codes (inclusion codes, Supplementary Table 12) and medications that can be used in the context of PD (inclusion medications, Supplementary Table 13). Then, for each PD case, a pool of candidate controls were identified based on exact matching of gender (self-reported), year of birth, race/ethnicity (self-reported), and the presence or absence of hypertension (ICD-10 code: I10) and type 2 diabetes mellitus (ICD-10 codes: E11, E11.01, E11.618, E11.620, E11.621, E11.622, E11.628, E11.630, E11.638, E11.641, E11.649, E11.65, E11.69, E11.8, E11.9). The pool of candidate controls for each PD case was further refined based on the timing of plasma donations, to match storage duration, and on donation frequency, to account for the impact of repeated plasma donation on the proteome. This refinement used two metrics: the temporal overlap of case and control donation windows and their donation frequencies. From these, we derived an overlap ratio and a frequency ratio, and ranked candidate controls by the product of these ratios to prioritize those with aligned sampling windows and similar donation frequency. Final Chronos Twin pairs were confirmed by visual inspection of donation plots. When no suitable match shared the exact year of birth, candidates within ±2 years were considered.

### Sample Selection, Processing, and Handling

We applied a rule-based framework to ensure consistent temporal resolution across donors. When only one or two samples were available, all were included. For donors with three or more samples, selection was based on donation span. Donors with ≤1 year of donations and ≥ 3 samples contributed three samples corresponding to the first, middle, and last draws. Donors with ≥ 1 year and ≤ 3 years of donations contributed at least four samples, including the first and last and two additional draws spaced at least six months from both. Donors with ≥ 3 years of donations contributed to ≥ 5 samples, comprising the first and last, two samples spaced approximately six months from the first and last, and additional samples spaced roughly one year from other selected draws. The same rules were applied to Chronos Twins to ensure comparable temporal resolution and sampling structure between each case and its matched control. Selected samples were retrieved from the long-term storage facility (Benson, North Carolina, USA), packed, and shipped frozen on dry ice to Alkahest, Inc. (San Carlos, California, USA). Upon receipt, all samples were immediately transferred and stored in -80°C freezers. Samples were thawed in a controlled manner, and a Hamilton automated liquid handling system was utilized to precisely aliquot the samples into volumes required per each proteomics vendor’s specifications. Sample aliquots were promptly returned to -80°C storage until required for shipment. Sample aliquots were fully randomized across plates and vendors, then shipped frozen on dry ice to each proteomics vendor for analysis. Plate load order was not controlled across platforms.

### Plasma Proteomic Profiling

To identify PD-associated proteomic signatures, we used a multi-platform strategy combining affinity-based profiling and mass spectrometry. Parallel analysis across all platforms generated a dataset of more than 25,000 features, enabling broad proteome coverage and robust cross-platform and external validation. All analyses were performed blinded to case-control status.

### SomaScan Assay

We used the SomaScan 11k Assay v5.0 (SomaLogic Inc., Boulder, CO, USA), a highly multiplexed, high-throughput, aptamer-based proteomics platform that measures 10,776 unique human protein analytes. It uses SOMAmer® (“Slow Off-rate Modified Aptamer”) reagents, which are single-stranded DNA molecules with chemical modifications that enhance protein-specific binding specificity and affinity. Each SOMAmer reagent adopts a defined three-dimensional structure and binds to proteins in their native folded conformations. SOMAmers are developed using SELEX (Systematic Evolution of Ligands by Exponential Enrichment) technology. During the assay, protein binding sites are converted into corresponding SOMAmer concentrations, which are quantified via microarray-based detection. Data normalization employs hybridization controls to correct for variance in hybridization, washing, and scanning steps. All analyses were performed using hybNorm.medNormInt.plateScale.calibrate.anmlQC.qcCheck.anmlSMP.adat file that includes ANML normalization. Results are reported as relative fluorescent units (RFU) for relative proteoform abundance. Log2 transformation is applied for downstream quality control and modeling. This dataset is referred as Somalogic-11k in the manuscript.

### Olink Explore HT

The Olink Explore HT platform measures 5,420 unique human proteins using Proximity Extension Assay (PEA) technology. PEA is an advanced multiplexed immunoassay designed to minimize cross-reactivity, a limitation of conventional immunoassays. Each antibody pair is conjugated to unique oligonucleotide sequences that hybridize only when both antibodies bind to the same target protein. This proximity-dependent hybridization enables DNA extension, creating unique reporter sequences that are subsequently amplified and quantified by next-generation sequencing (NGS). This design eliminates cross-reactivity because only correctly paired oligos generate an amplicon. Data are normalized using internal extension controls and inter-plate median normalization, and results are reported as NPX (Normalized Protein eXpression) values on a log2 scale for relative protein quantification. This dataset is referred as Olink-HT-5k in the manuscript.

### NULISAseq™ CNS Disease Panel 120

Alamar Biosciences’ NULISAseq™ CNS Disease Panel 120 measures 127 unique human proteins or peptides using NULISA (NUcleic acid Linked Immune-Sandwich Assay) technology. This method employs two DNA-antibody conjugates that simultaneously bind to the target protein, forming a sandwich complex. The conjugated DNA strands are then ligated, followed by purification steps to reduce background noise. Quantification is performed using quantitative PCR (qPCR) for low-plex panels or next-generation sequencing (NGS) for high-plex panels. NULISAseq data are normalized using internal controls and inter-plate median normalization. Results are reported as NULISA Protein Quantification (NPQ) units on a log2 scale for relative protein abundance. This dataset is referred as AlamarBio-CNS-120 in the manuscript.

### Discovery Mass Spectrometry

Untargeted proteomic profiling was performed using the Biognosys TrueDiscovery™ platform with integrated P2 Plasma Enrichment. Plasma samples were processed on a KingFisher Flex (Thermo Scientific) according to Biognosys’ Standard Operating Procedures, consisting of protein enrichment using proprietary beads for P2, reduction, alkylation, and digestion to peptides using trypsin (Promega, 1:50 protease to total protein ratio) and Lys-C (Fujifilm Wako Chemicals, 1:200 protease to total protein ratio) overnight at 37 °C. Clean-up for mass spectrometry was performed using an Oasis HLB μElution Plate (30 μm) (Waters) according to the manufacturer’s instructions. Peptides were dried down using a SpeedVac system and dissolved in LC solvent A (1% acetonitrile/0.1% formic acid (FA) in water) containing Biognosys’ iRT-peptide mix for retention time calibration. Peptide concentrations in mass spectrometry-ready samples were measured using a Micro BCA assay (Pierce, Thermo Scientific). 800 ng of peptides were loaded on EvoTip according to the manufacturer’s instructions. Samples were prepared separated using the Whisper Zoom 40 SPD method on an Evosep One using an IonOpticks Aurora® Elite™ 15 cm x 75 µm C18 UHPLC column. Data were acquired in diaPASEF® mode using one full-range MS1 scan (100-1,700 m/z) with an ion mobility window of 0.85 – 1.45 1/k0 across 8 PASEF ramps. In-batch calibration was enabled using the Tuning Mix ES-TOF CCS reference list as the calibration template. Data was processed in Spectronaut® 19.3. Searches were performed against a UniProt FASTA database (Homo sapiens, 2024-07-01), with sparse setting, allowing 2 missed cleavages, fixed modification (carbamidomethylation), and variable modifications (N-term acetylation, methionine oxidation, and ammonia loss). Peptide and protein identifications were filtered at a 1% false discovery rate (FDR). Missing or sub-threshold values were imputed based on background noise distribution. Protein quantities were calculated with Quant2.0 using default settings, including cross-run normalization by local regression. This dataset is referred as discovery MS in the manuscript.

### Statistical Analysis

All analytical dataset preparations, visualizations, and statistical analyses were performed using R version 4.4.3 or higher. Unless otherwise specified, an alpha of 0.05 and two-sided tests were used for significance testing, unless otherwise stated. To adjust for multiple comparisons, p-values were corrected using the Benjamini-Hochberg FDR method and reported as *q*-values. Effect sizes including Cohen’s d and spearman correlation (rho) were reported besides p-values and q-values when appropriate. Cohen’s d was calculated using *t_to_d* function from *effectsize* v1.0.1 R package after linear mixed modeling, and rho was calculated using *cor* function from *stats* R package or *cor.test* function from *stats* R package when p-values were also reported.

In this study, we use proteoform to mean a platform specific measured analyte (assay) that may map to one or more UniProt proteins, whereas protein refers to the UniProt ID level after mapping assays to proteins (splitting multi protein annotations and collapsing multiple assays per protein when needed).

### Descriptive Summaries

Baseline characteristics including demographic variables, donation patterns, and variables derived from claims and medication data were summarized. For continuous variables, descriptive statistics included the number of observations, number of missing values, mean, median, minimum, maximum, and standard deviation (SD). For categorical variables, frequency counts and percentages were reported. Spearman’s correlation was used to assess univariate associations between continuous variables. Between-group comparisons were performed using Chi-square tests or Fisher’s exact tests for categorical variables and two sample t tests or Wilcoxon rank sum tests for continuous variables.

### Comorbidities Analysis and Comparison to US Population-Level PD Cohort

To better understand the clinical features of PD in our cohort, identify conditions that co-occur or precede its onset and compare their prevalence against population-level data, we performed a comorbidity enrichment analysis in the Chronos-PD cohort (n=348) with matched controls (n=348) and in an established US population-level PD cohort (n=216,336) with matched controls (n= 216,336). The US population-level PD cohort was derived from the full RWD database, which covers more than 400 million individuals and is not restricted to plasma donors. It was defined using the same criteria as our *established PD cohort* (see ‘Identification of Parkinson’s Disease Cohort’), and controls were selected using the same approach, except without matching on sample characteristics. Comorbidities were defined as ICD-10 code categories, with individual codes being collapsed down to three digits to form broader categories. Events were categorized as “pre-onset” or “post-onset” depending on whether the date of a comorbidity was prior to or after the estimated date of PD onset. For each Chronos Twin pair, the onset date from the PD case was applied to the control to ensure temporal alignment.

Separate design matrices were constructed for pre-onset and post-onset windows. For each matrix, subjects formed the rows and ICD-10 categories the columns; a cell was coded 1 if the subject had at least 1 record of that category within the respective time window, and 0 otherwise. Duplicate rows (multiple events per subject–code) were collapsed by taking the per-subject maximum. Each matrix was then augmented with two covariates: year of birth (continuous) and gender (binary). The dependent variable “group” was encoded 1 = case, 0 = control.

Comorbidity enrichment was quantified using odds ratios (ORs) derived from logistic regression models:

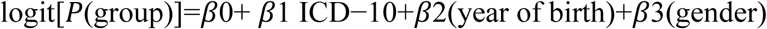

Parameter estimation used the mean-bias-reduced maximum likelihood method (*brglmFit()* function from the *brglm2* v0.9.1 R package) to avoid infinite estimates arising from complete separation. For each ICD-10 category, we report the odds ratio (OR = exp β₁), Wald 95 % confidence interval (CI, profiled on the bias-reduced scale) and two-sided p-value. False discovery rate (FDR) was controlled independently for pre-onset and post-onset windows using the Benjamini-Hochberg correction.

### Proteomics QC

To ensure proteomics data integrity and platform reliability, we first assessed proteomic data quality by calculating technical coefficients of variation (CVs) from internal replicate samples (n=3 samples, each measured in triplicates) and by determining the percentage of samples below the limit of detection (LOD). Samples that failed Olink quality control (n = 2) or carried warnings (n = 23) were removed. Plate and batch effects were evaluated using multiple approaches, including visualization of their impact on the first two principal components (PC1 and PC2) from Principal Component Analysis (PCA), inspection of median protein values per plate, and random plate-effect models. Based on these assessments, 5 plates from AlamarBio-CNS-120 and 2 plates from discovery MS were rerun. Additional median based plate normalization was applied to the discovery MS data to correct for pronounced plate effects.

Outlier detection was performed using two complementary criteria: (1) values exceeding six median absolute deviations (MAD) from the first two PCA components and (2) values exceeding five standard deviations (SD) from the mean of median sample signals. Samples meeting both criteria were removed, resulting in the exclusion of 11 Biognosys MS samples. No samples were removed due to protein values below LOD, and no proteins or assays were excluded for poor technical CVs or high proportions of samples below LOD.

### Identification and justification of confounding factors for statistical modeling

Proteomic levels and their longitudinal changes can be strongly influenced by biological and technical covariates. When these covariates differ significantly between PD and control donors, they act as confounders and must be accounted for to avoid biased results. Known potential confounders including age, gender, race, health status, donation patterns, and storage duration were addressed during the cohort design stage through Chronos Twin matching.

Additional potential confounders were explored using dimensionality reduction techniques such as PCA (*prcomp()* function from *base* R package), tSNE (*tsne()* function from *Rtsne* v0.17 R package), and UMAP (*umap()* function from *uwot* v0.2.4 R package). All three methods revealed two distinct clusters in Somalogic-11k, Olink-HT-5k, and AlamarBio-CNS-120. To characterize this latent factor, PC1 loadings from Somalogic-11k were extracted, and the top 500 proteins contributing most to PC1 variance were selected. Hierarchical clustering (*hclust()* function from *base* R package) was then performed using Euclidean distance and Ward’s minimum variance method (ward.D2), which minimizes within-cluster variance. This analysis defined two clusters (PC_cluster), which were included as covariates in downstream models. Internal analyses indicate that these clusters align with fasting status, suggesting the latent factor reflects fasting-related physiological variation. Group differences were examined, and the impact of these factors on proteomic variation was quantified using the *relaimpo* v2.2.7 R package. Based on these analyses, we confirmed the following as key covariates for modeling proteomic data: age at baseline (defined as the age at the first sample selected for proteomics measurement in this study), gender, race, recent donation status defined as any donation within 30 days, and the data-driven PC_cluster variable. Technical effects such as center and plate variability were also detected for certain assays and platforms and were adjusted in the linear mixed models described below.

### Detection of proteomic changes over time during PD progression

To identify proteomic changes significantly associated with PD progression, we applied a two-step modeling strategy. First, a linear mixed model (referred as screen model) was used to detect PD-related signals under the assumption of a linear relationship between protein levels and years since estimated PD onset. Second, two non-linear mixed models were applied to identify proteins with non-linear trajectories and their inflection points.

For the screen model, we used the *lmer()* function from the *lmerTest* R package, allowing flexible random-effects structures, and selected the best-fitting model for each protein based on the Bayesian Information Criterion (BIC). Two fixed-effect specifications were evaluated: one including age at baseline, gender, PC_cluster, and recent donations, and a second adding race as an additional covariate. Four random-effect structures were considered: a donor-level random intercept; random intercepts at both donor and center levels; a random intercept with a correlated random slope; and a random intercept with an uncorrelated random slope. These models allow individual trajectories to deviate from the group mean through random intercepts and slopes. Their purpose was to identify PD-associated signals reflected either in absolute protein levels or in differential rates of change between cases and controls. Because the Biognosys Discovery MS data showed substantial plate effects, plate ID was included as a random effect in these models.

For significant signals (q < 0.05), two non-linear mixed models were applied. A linear spline mixed model was used to identify the earliest divergence time between PD and control groups when it outperformed the best linear mixed model based on BIC (BIC difference > 2). Using *lmerTest::lmer*, we tested thirteen candidate knot positions spanning from 7 years before to 5 years after estimated PD onset. Assays with smaller BIC values and at least one significant slope (p < 0.05) were retained for further evaluation. A natural cubic spline mixed model was also used to determine the time point corresponding to the fastest rate of change for each protein when it provided a better fit than the linear mixed model (BIC difference > 2). This model, implemented with lmerTest::lmer, was tested with two to four knots using the best selected mixed-model structure, and assays with smaller BIC values were retained for downstream analysis. Model assumptions, including residual normality and homogeneity of variance, were examined to ensure agreement between model and data for identified PD proteins.

### Treatment exposure effect on proteomics profiles

Post-onset samples (n=435) were classified according to the presence or absence of levodopa-containing medication (Supplementary Table 15) prior to sample collection.

After further filtering, 113 samples were selected for the analysis and divided into 2 groups: levodopa+ samples with levodopa prescription one month prior to sample collection date (levodopa+, 29 individuals, 55 samples) and levodopa- samples with other ongoing medication records around the samples and without any levodopa prescription in one year prior to sample collection date (levodopa-, 25 individuals, 58 samples). To minimize potential misclassification due to incomplete medication records, samples lacking medication data for both the 3 months before and 3 months after the donation date were excluded. Prior to testing for levodopa-associated proteomic changes, we evaluated whether the groups differed in relevant non-protein covariates, including PD onset, gender, birth year, race, diabetes, hypertension, cluster, and recent plasma donation frequency. To assess potential levodopa-associated effects among the top PD protein markers, we compared protein levels between the groups of samples using linear mixed-effects models. Protein abundance was modeled with medication group as the primary fixed effect of interest and years since PD onset, gender, age at baseline, recent plasma donation history, and PC_cluster included as covariates. A subject-specific random intercept was used to account for repeated measurements within individuals.

### Identification of PD risk biomarkers using joint modeling of longitudinal proteomic data and survival data

Identifying biomarkers associated with PD risk is essential for early detection and stratification. Traditional linear and non-linear mixed models rely on “years since onset,” a variable inferred from the first G20 diagnosis or PD medication claim and therefore susceptible to real-world data limitations. For controls, onset timing is inherited from matched PD donors under the digital-twin assumption and carries the same uncertainties. To better capture the dynamic relationship between protein levels and PD risk, we applied joint modeling^39^ of longitudinal proteomic data and time-to-PD survival to the top Chronos-PD signals identified by linear mixed models. This approach was applied to proteomic datapoints collected before the estimated date of PD onset, and results were compared with those obtained from the previously described linear and non-linear mixed models. The time-to-PD survival table was generated as age of the first record in claims table, age to PD (event) or censored with claims table, and modeled using a Cox proportional hazards regression model (*coxph()* function from *survival* v.3.8-3 R package) adjusted with gender. Longitudinal proteomic measurements were modeled over age at sample collection using a linear mixed model with fixed effect of gender, PC_cluster, recent donations and random intercept and random slope. The association between biomarker levels and the risk of PD onset was estimated by Bayesian joint longitudinal-survival model implemented with the *jm()* function from *JMbayes2* package. Parameter estimation was performed via Markov chain Monte Carlo (30,000 iterations with a 3000-iteration burn-in across 4 chains, with thinning applied every 2 iterations). Priors for model parameters were derived from the separately fitted longitudinal and survival submodels. Convergence diagnostics were performed using the Gelman-Rubin R-hat (rhat) statistic, with value below 1.05 interpreted as evidence of satisfactory convergence across chains. The identified proteins were ranked based on the estimated risk of PD onset for 0.1 unit change in protein levels and associated p-values. This joint modeling approach accounts for informative dropout in claims data and provides direct estimates of PD-onset risk per unit change in biomarker levels. Because it excludes proteomic measurements collected after onset, the analysis was limited to 2,095 samples from 240 PD donors and 328 controls.

Chronic comorbidities may confound the identification of PD-risk biomarkers or help support their relevance. To distinguish biomarker associated with PD comorbidities from those linked to other chronic conditions, we applied joint modeling to capture the dynamic relationship between comorbidity risk and longitudinal protein trajectories. Chronic comorbidities were identified using the Chronic Condition Indicator Refined (CCIR) for ICD-10-CM, yielding 12,769 chronic ICD-10 codes out of 75,238 total codes. For analysis, we aggregated diagnoses to their 3-characters ICD-10 groups, resulting in 451 unique categories, of which 449 remained after removing G20 (Parkinson’s disease) and G21 (secondary parkinsonism) codes. We then evaluated the association between each of the 449 chronic comorbidities and PD using odds ratios (*fisher.test()* function from *base* R package) with Benjamini–Hochberg correction (*p.adjust()* function from *base* R package). Fifty comorbidities met significance (q < 0.05) and were designated “PD-chronic comorbidities,” while 52 additional comorbidities with q ≥ 0.05 and at least 30 individuals were classified as “other chronic conditions”. Joint modeling for these comorbidities was performed as described above.

### Biomarker trajectory groups

We classified PD associated biomarker trajectories by integrating linear mixed models, non-linear mixed models (linear spline and natural cubic spline), and joint modeling. We excluded six proteins with strong medication effects (DDC, NAA10, OIP5, ZC3H12C, CYLD, and TUBA1A), defined as |Cohen’s d| > 0.8 with p < 0.05. All biomarkers considered for trajectory grouping were required to be significant in the mixed model that included all 2,609 samples measured in this study (q-value < 0.05). Non-linear models were retained when they improved fit over the corresponding linear model (ΔBIC > 2). Biomarkers were then assigned to three trajectory groups: (i) Early linear change for biomarkers that were also significant in linear mixed models that included pre-onset samples only (n=1748, q-value < 0.05) and/or in PD risk joint models (n=2,095, p < 0.01 with rhat < 1.05) (ii) Inflection years before onset for biomarkers with evidence of pre-onset non linearity (ΔBIC > 2 and an inflection time at/or before PD estimated onset) and a significant group difference in the first spline segment (first slope, p < 0.05) (iii) Late linear and non-linear for biomarkers that did not meet criteria for groups (i) or (ii), including those with post linear rate of change driven by post PD onset signals (q-value < 0.05) and/or better fitting non-linear trajectories with late divergence, reflected by a significant second spline segment or post onset effect (ΔBIC > 2 with p < 0.05)

### Pathway enrichment analysis of top PD signals

We queried pathway enrichment databases to interpret the biological significance of the PD identified proteins. Several proteoforms measured in this study were mapped to multiple UniProt IDs in the annotation provided by proteomics vendors, which can arise from factors such as measuring protein complexes. To enforce a single protein assignment per assay within each platform, we applied a two-step disambiguation strategy. First, we identified the “core” set of assays which are associated with only a single UniProt ID, each. Then, for assays with multiple possible associations, we picked the association already covered by this “core” set. This practice makes the analysis conservative: if a signal is detected by multiple assays, we try to explain the origin of that signal with the smallest possible set of proteins detected. Second, if the rule above did not produce an unambiguous selection for an assay, we summarized the number of biological associations of the possible associated proteins within Gene Ontology (GO)^40^, Kyoto Encyclopedia of Genes and Genomes (KEGG)^41^ and Reactome^42^ databases. The UniProt ID with the most known biological annotations was picked to facilitate descriptive power of the analysis. In cases where the above methods failed, we ranked the mapped UniProt IDs alphabetically and picked the first association for the analyte. Then, results from the three affinity platforms were combined (Discovery Mass Spec was not included due to the absence of significant PD signals). Their unified coverage (11,260 Uniprot IDs) was used as a background for enrichment analysis. For proteins measured by multiple assays from different platforms the lowest p-value was selected.

Then, within each mixed model analysis (pre-diagnosis, post-diagnosis, all samples), upregulated and downregulated proteins were ranked separately based on p value. Ties were resolved by absolute Cohen’s D. Those rankings formed the basis of Sliding Enrichment Pathway Analysis (SEPA^43^). From each ranked list, 50 interlocking protein sets were established, consisting of the top n = 10, 20, … , 500 proteins (SEPA depths). Within these sets, Overrepresentation Analysis (ORA) was performed to detect enrichment of GO, Reactome and KEGG terms. On results, FDR correction was applied in a per-set manner. GO Biological Process, GO Molecular Function, GO Cellular Component, KEGG and Reactome terms were handled separately. An enrichment was accepted if the term overrepresentation exceeded the FDR-corrected significance cutoff (q-value < 0.05) in at least 3 consecutive top sets. A comprehensive summary of SEPA results can be browsed in Supplementary Table 6 detailing the most significant enrichment and the first SEPA depth passing the filtering criteria for each pathway in each condition.

### Visualization of enriched pathways and associated proteins

To obtain a comprehensive yet compact visualization of enriched pathways and their links with proteins, we first selected *GO Biological Process, Reactome* and *KEGG* pathways that met a stringent SEPA significance criterion in at least one of the three analyses (all samples, pre-onset, post-onset). Pathways were retained if they showed FDR < 0.01 in three or more consecutive SEPA depths among the top 10-150 proteins (evaluated in steps of 10 proteins), had at least three associated proteins with substantial change (fold-change ≥ 2 in the tested direction) and contained between 30 and 2000 quantified proteins in the background. For each pathway, we determined the dominant direction of regulation across cohorts and classified it as upregulated, downregulated, or bi-directional (mixed behavior). Pathways were scored based on their peak significance value on the investigated SEPA range and the first SEPA depth they appeared as confirmed hits using the formula:

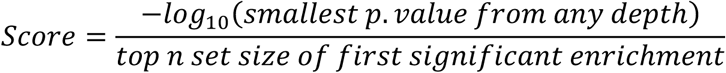

Then, within each of the six analysis and directionality categories, pathways were ranked based on their score.

Upregulated, downregulated and bi-directional pathways were then clustered separately, based on overlap of their associated gene sets: a pathway–gene incidence matrix (GO, KEGG and Reactome) was used to compute pairwise Jaccard similarities, and clusters were obtained with the *Louvain* method implemented in the *simplifyEnrichment* R package. For each cluster, the highest-ranking pathway across the three analyses (with ties broken with preference to the smallest number of associated proteins in the measured background) was chosen as the representative cluster label in the heatmap. For the final visualization, each heatmap pixel represents the strongest evidence for enrichment within a cluster at a given cohort and SEPA depth, defined as the minimum FDR among all pathways in that cluster at that depth. Values are plotted as signed -log10(FDR), with positive values indicating upregulation and negative values indicating downregulation; non-significant values (FDR ≥ 0.05) are set to zero.

To visualize the network of top pathways and associated proteins, we focused on four SEPA analyses (pre-onset upregulated, pre-onset downregulated, post-onset upregulated, post-onset downregulated). For each analysis, we first identified pathways that met the SEPA significance filter detailed above. The union of the top five pathways from each analysis was defined as the term set of the network. As the protein set, we used all proteins present among the top 20 proteins in any of the four analyses and removed proteins not associated to any selected pathway. For visual simplification, proteins with identical pathway neighborhoods and presence in identical analysis top 20 sets were collapsed to single nodes (labeled by concatenated names in alphabetical order).

In the final visualization (generated with the *igraph* and *visNetwork* R packages), pathway nodes are rectangles and protein nodes are circles; color encodes pre-onset and post-onset regulation.

### Protein ratio analysis

Ratios are valuable biomarkers in neurodegenerative diseases because they normalize individual variability and highlight relative changes that can be more biologically informative than absolute levels. In PD, ratios capture interactions across pathways and help mitigate batch effects and inter-individual variability by relying on relative rather than absolute measurements. We generated pairwise protein ratios from all assays across three affinity platforms (Somalogic-11k, Olink-HT-5k, and AlamarBio-CNS-120), resulting in over 131 million within-platform and cross-platform pairs. We evaluated the predictive performance of these ratios across multiple time windows relative to PD onset, including six 2-year sliding windows [-8, -6), [-7, -5), [-6, -4), [-5, -3), [-4, - 2), [-3, -1). For each window, one sample per matched PD-control pair was used; when multiple samples were available for a subject, the middle sample was selected. At each time window, protein ratios were generated, and samples were split into training (2/3) and testing datasets (1/3) and 3-fold cross validation (CV) was performed. Logistic regression (*glm()* function from *base* R package) adjusted with age and gender were used to build prediction model in training datasets and evaluated in testing datasets. AUCs, accuracy, sensitivity, specificity of mean of the 3-fold CV were recorded. The discovery step aimed to identify ratios with AUC > 0.7 in the close-to-onset window [-3, -1), and the subsequent prioritization step focused on ratios that also achieved AUC > 0.6 in 2+ additional pre-onset two-year windows. This approach enriched for candidate biomarkers with potential three-year risk or diagnostic value. As a comparison to the ratio analysis, we repeated the [-3, -1) analysis for single proteins and built a multivariate classification model using XGBoost (*xgb.train()* function from the *xgboost* v1.7.8.1 R package) using a three-fold cross-validation framework with ten repeated fold redistributions. Elastic-net logistic regression (α = 0.5) was also implemented using the *glmnet* v4.1.8 R package. The regularization parameter λ was selected by 5-fold cross-validation maximizing AUC on the train set, and predictions were obtained from the model at λ_min for the test set.

### Comparison between Chronos-PD signals and literature-reported PD signals

Results from our analysis were benchmarked against 17 cohorts, spanning 36 analyses and approximately 4,000 research participants with PD (Supplementary Tables 3 and 16). Significance thresholds were taken directly from each study (Supplementary Table 16) to ensure comparisons reflected cohort-specific criteria and avoided imposing a uniform threshold across heterogeneous datasets. Literature-based PD signals spanned plasma, CSF, and urine across multiple proteomic platforms. Because these fluids capture complementary biological processes, integrating them increases biomarker confidence and minimizes fluid-specific artifacts. PD signals detected in our study were compared at the protein level using UniProt IDs, and consistency in effect sizes and directionality was assessed with Spearman correlation coefficients

### Validation of molecular signals in external PD cohorts

To validate single proteins Chronos-PD signals and protein ratios, we assessed their consistency across three external PD cohorts: the PPMI-p9000 project, three cohorts from the Global Neurodegeneration Proteomics Consortium (GNPC V1 harmonized dataset), and the UK Biobank cohort. GNPC and PPMI represent clinically-phenotyped cohorts composed primarily of individuals with prevalent PD, whereas UK Biobank is a population-based study that includes mostly incident PD cases.

PPMI-p9000 project used here was downloaded on 15-Sep-2025. Proteins were measured by Olink 1.5k platform and measurements are originally from two studies (projects 196 and 222), including a total of 78 PD patients and 130 controls (patients labelled as prodromal patients were not included), followed for up to 15 visits over as many as 8 years. For each subject, the most recent sample was used for validation. Three cohorts in GNPC (cohorts C, J and L) were selected based on the criteria that there were >50 samples in each of PD and control groups after matching and cohort feature characterization. Cohort C and L were cross-sectional datasets and contained Somalogic-7k measurements from plasma EDTA samples. Cohort J was a longitudinal dataset with both plasma and CSF samples. For each subject, only the most recent plasma EDTA sample for validation. For PPMI-p9000 and GNPC cohorts, quality control was performed to check for outliers, potential confounders and covariates (see above). No outliers were detected in PPMI-p9000 proteomic dataset, and 51 (2.55%), 21 (4.98%), 6 (1.4%) outliers were removed for Cohort C, J and L, respectively. Significant differences in age, gender, and cognitive function were observed between PD cases and controls in the GNPC cohorts. To mitigate these confounding effects, propensity score matching (1:2 or 1:1) was performed on age, sex, and/or Alzheimer’s disease status using *matchit()* function from *MatchIt* R package based on nearest mahalanobis distance. This resulted in 126 PD cases and 252 controls in cohort C, 99 PD cases and 99 controls in cohort J, and 66 PD cases and 132 controls in cohort L. Two additional GNPC cohorts (F and Q) also included PD cases; however, QC analyses revealed strong latent factors in cohort F and demographic disparities in cohort Q, including a small sample size (n = 57) and lower education levels relative to other cohorts, resulting in limited detection power. These cohorts require further investigation using information beyond what GNPC provides, so their results are reported only in Supplementary Fig. 4.

UK biobank data used here was downloaded on 22-Nov-2024. UKB-PD individuals with PD were defined based on having a G20 ICD-10 code in Data-Field 41270. A cohort of non-PD participants was defined as those participants with no G20 ICD-10 code in either Data-Field 41270, Data-Field 41202 or Data-Field 131022. For each participant in the UKB-PD cohort, a single matching control was identified from the control pool using nearest neighbor matching, as performed using the MatchIt R package with default arguments in R. Variables included in the matching formula were: year of birth (Data-Field 34), sex (Data-Field 31), Type 2 Diabetes status (presence of an E11 ICD10 code in Data-Field 41270), hypertension status (presence of an I10 ICD-10 code in Data-Field 41270), Townsend deprivation index (TDI) at recruitment (Data-Field 22189) and a binary variable indicating if the participant had at least 1 protein measured for instance 0 (Data-Field 30900). Exact matching was required for all variables, except TDI. The date of PD onset was defined using the Date G20 first reported field (Data-Field 131022). Years since PD onset was calculated between the Date of attending assessment centre (Data-Field 53) and the date of PD onset, and two sliding window datasets were generated as described in Chronos-PD phase 1 dataset for [-9,-5) and [-5,-1) years before first G20.

Identified proteins from Chronos-PD signals or each pair of protein ratios were mapped to the external datasets based on UniProtIDs or gene symbols when UniProtIDs were not available. To validate single proteins Chronos-PD signals, linear regressions adjusted with age (or YOB) and gender were applied for UKB and GNPC cohorts (*lm()* function from *stats* R package). Linear mixed models with random intercept and fixed models with group*event_id, age at the first visit, gender were used for PPMI-p9000. To validate protein ratios, logistic regressions adjusted with age and sex were conducted. Estimated coefficient, SE, p-value, OR and 95% CI were recorded.

## Data availability

An interactive Shiny application for exploring the Chronos-PD dataset and results is accessible at: https://chronos-pd-proteomics-explorer.share.connect.posit.cloud/

The Chronos-PD datasets generated and analyzed during the Chronos-PD Phase 1 study will be made available to the research community upon publication. The portal will provide access to the data and support interactive exploration and querying of proteomic and RWD. Shared data will include a comprehensive data dictionary, privacy remediation guidance, and standardized formats aligned with FAIR (Findability, Accessibility, Interoperability, and Reusability) principles. Access to the portal will be subject to data use agreement.

Individual-level UK Biobank data are available under restricted access due to participant confidentiality and data privacy regulations. Researchers can apply for access through the UK Biobank Access Management System: https://www.ukbiobank.ac.uk/enable-your-research/apply-for-access. Access is granted to bona fide researchers for health-related research, following a formal application and approval process. The UKB data used in this study were generated by the UK Biobank Pharma Proteomics Project (UKB-PPP), obtained under the UKB project 82395. The data used in the preparation of this article was obtained on 11/22/2024.

The GNPC V1 HDS is accessible by the broader research community as a shared, global resource via the AD Data Initiative’s data platform, the AD Workbench (for approved use/users) as of July 15th, 2025.

PPMI Data used in the preparation of this article was obtained on 09/15/2025 from the Parkinson’s Progression Markers Initiative (PPMI) database (www.ppmi-info.org/access-dataspecimens/download-data), RRID:SCR_006431. For up-to-date information on the study, visit www.ppmi-info.org. PPMI – a public-private partnership – is funded by the Michael J. Fox Foundation for Parkinson’s Research and funding partners, including 4D Pharma, Abbvie, AcureX, Allergan, Amathus Therapeutics, Aligning Science Across Parkinson’s, AskBio, Avid Radiopharmaceuticals, BIAL, BioArctic, Biogen, Biohaven, BioLegend, BlueRock Therapeutics,

Bristol-Myers Squibb, Calico Labs, Capsida Biotherapeutics, Celgene, Cerevel Therapeutics, Coave Therapeutics, DaCapo Brainscience, Denali, Edmond J. Safra Foundation, Eli Lilly, Gain Therapeutics, GE HealthCare, Genentech, GSK, Golub Capital, Handl Therapeutics, Insitro, Jazz Pharmaceuticals, Johnson & Johnson Innovative Medicine, Lundbeck, Merck, Meso Scale Discovery, Mission Therapeutics, Neurocrine Biosciences, Neuron23, Neuropore, Pfizer, Piramal, Prevail Therapeutics, Roche, Sanofi, Servier, Sun Pharma Advanced Research Company, Takeda, Teva, UCB, Vanqua Bio, Verily, Voyager Therapeutics, the Weston Family Foundation and Yumanity Therapeutics.

## Code availability

All code used in the analysis and figure generation for this study will be made available on GitHub at https://github.com/AlkahestInc/chronos_pd_phase_1_manuscript and will be provided to reviewers upon request.

## Acknowledgments

This research was supported by funding and oversight from the Michael J. Fox Foundation (MJFF-025254 and MJFF-024708). We thank Nicole Vagle and the team from the plasma logistics centers; Balazs Szoke for support with sample logistics; Sanket Rege, Yesenia Lopez, and Onkar Dhande for the specialist review of medical records; the engineering team for the data infrastructure; and Madison Kendrick, David Winters, and Tracey Horne for their support of this work, as well as the Grifols plasma donors. We also gratefully acknowledge the guidance of our Scientific Advisory Board and thank Sonya Dumanis, Andy Singleton, and Andy Conrad for their contributions as members of the Board.

## Author contributions

**Conceptualization and overall project supervision:** B.L. S.L. **Study design:** Oversight: B.L. C.F. I.K. with contributions from S.L. N.W-H. K.N. S.G. B.M. S.P. T.W-C. Case identification I.K. with support from S.G. B.F. A.T. Controls matching C.F. I.K. R.T. N.H. N.W-H. Samples selection C.F. Y.W. **Data integration:** Tokenization B.L. with support from S.L. Data streams aggregation K.D. R.Sa. S.L. I.K. N.H-W. Data cleaning I.K. N.H. B.F. N.W-H R.Sa. S.L. Software engineering and internal tooling K.D. R.Sa. S.L. N.W-H. R.T. **Sample logistics:** S.L. with support from K.D. B.L. **RWD:** Oversight I.K. analysis I.K. N.H. Preparation for joint modeling B.F. R.Su. **Proteomics:** Oversight. C.F. Quality control Y.W. Y.G. C.F. T.N. Linear mixed and spline models C.F. Y.W. Y.G. Platforms comparison: Y.W. Joint modeling Y.G. Y.W. C.F. Medication effect B.F. Pathways enrichment T.N. Ratios analysis. Y.G. N.W-H. External validation Y.W. J.G. Y.G. T.N. N.W-H. Shiny app YW. R.Sa. **Manuscript:** Drafting and figures creation B.L. K.K. C.F. I.K. B.F. N.H. N.H-W Y.W. Y.G. T.N. Edition: B.L. K.K. with support from I.K. C.F. All authors reviewed the manuscript. **Funding acquisition:** B.L. S.L. with support from K.N.

## Additional information

Correspondence should be addressed to Benoit Lehallier and Scott Lohr

## Peer review information

[placeholder]

---

**Supplementary Fig. 1:**
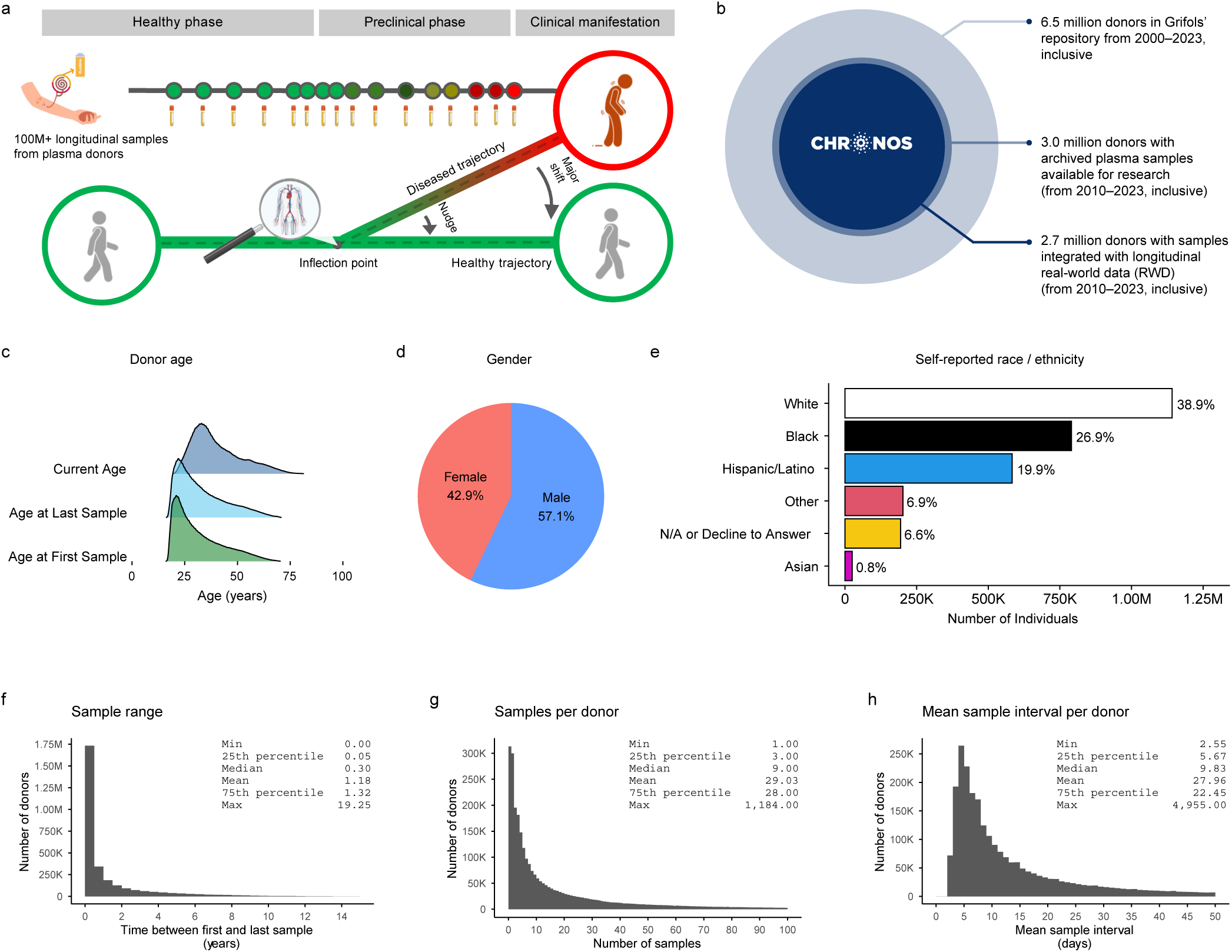
Characterization of the plasma donor registry used in this study. **a**, Conceptual overview of longitudinal plasma sampling across the disease continuum, illustrating divergence of healthy and disease-associated trajectories around an early inflection point. **b**, Overview of the plasma donor registry scale, indicating the progression from 6.5 million total plasma donors to the 3 million donors with biospecimens available for research, and the 2.7 million donors with integrated longitudinal medical data. **c-e**, Demographics of the 3 million donors from Grifols Registry with biospecimens available for research. Age distributions for donors at first available plasma sample, last available plasma sample, and current age (**c**), distribution of gender (**d**) and self-reported race / ethnicity (**e**). **f-h**, Additional descriptive metrics capturing the breadth, depth, and temporal resolution of plasma donations. X-axes are truncated to reduce the influence of long tails and improve visualization of the distributions.

**Supplementary Fig. 2:**
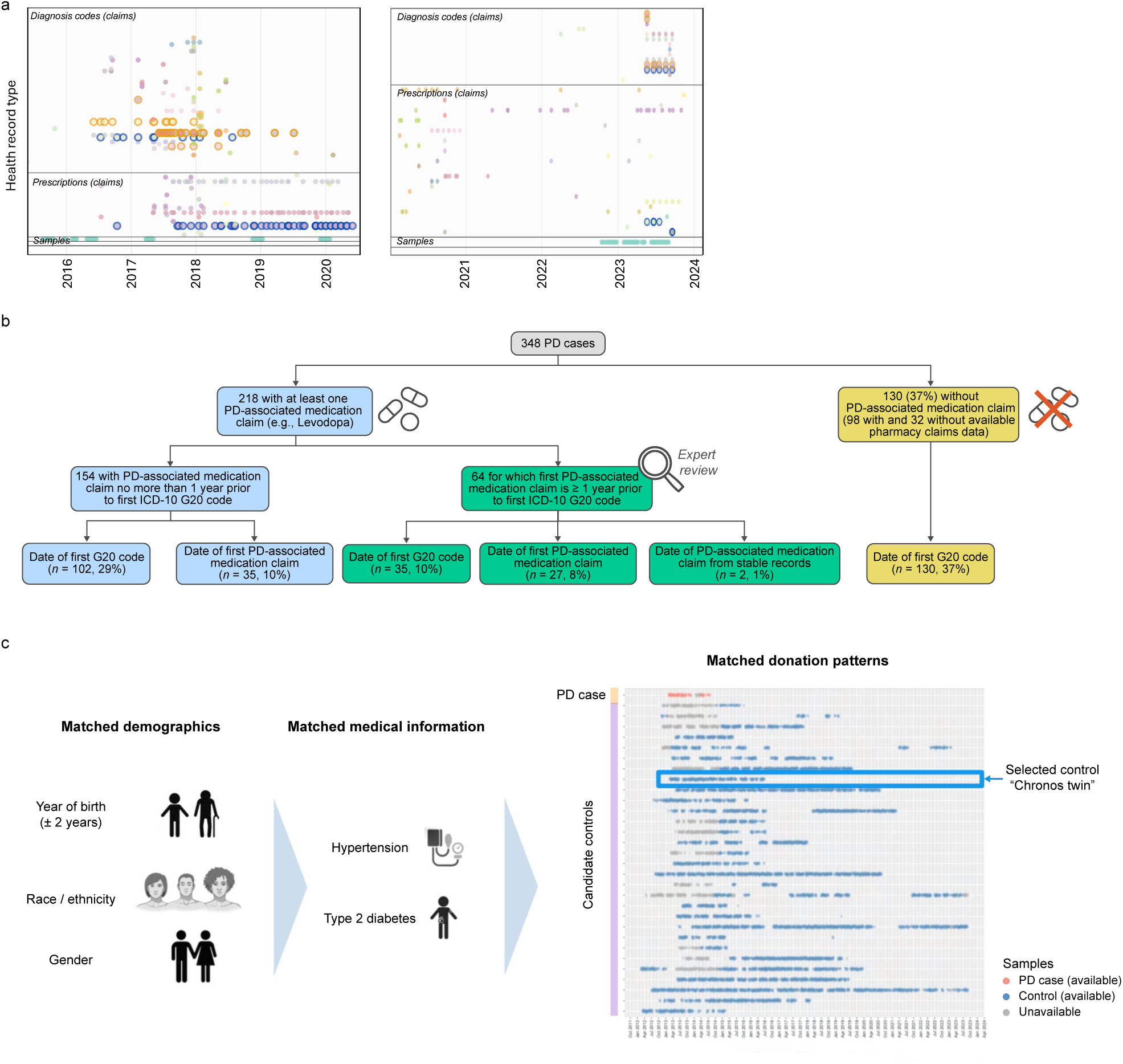
Cohort definition and controls matching. **a**, Representative examples of longitudinal health journeys for two plasma donors, with each dot representing a clinical encounter and colors indicating diagnosis codes, medications or samples; PD-related ICD-10 codes are highlighted. **b**, Decision tree for estimating PD onset date, integrating timelines of G20 ICD-10 codes and PD medication claims. **c**, Schematic representation of the matching process, based on demographic profiles (year of birth, race / ethnicity, and gender), clinical metadata (presence or absence of hypertension, type 2 diabetes), and timing / frequency of plasma donations.

**Supplementary Fig. 3:**
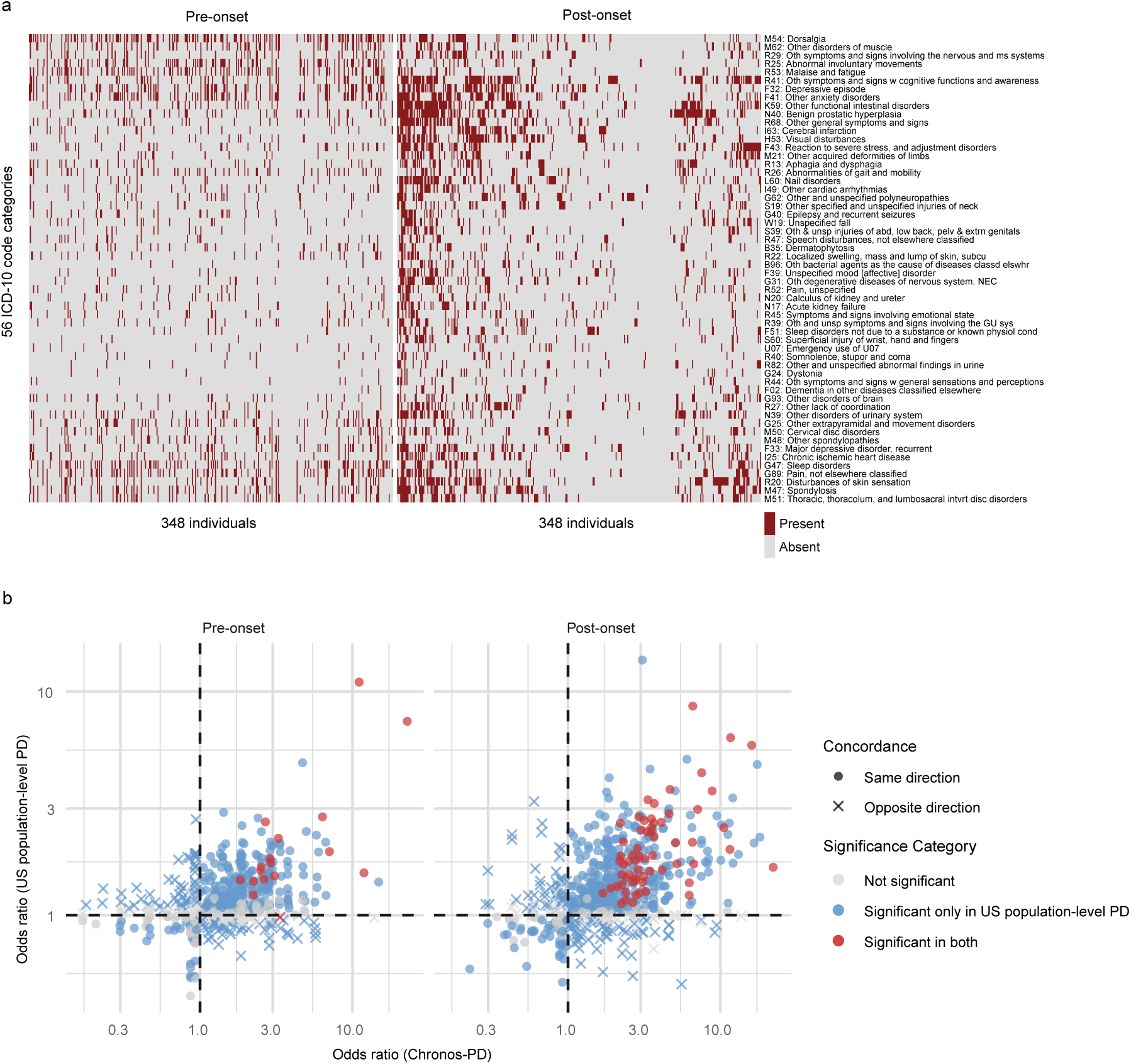
Comorbidities landscape across cohorts. **a**, Distribution of PD comorbidities in Chronos-PD cases pre-onset (left) and post-onset (right). **b**, Directional concordance of log odds ratios for comorbidities between Chronos-PD and US population-level PD cohort, pre- and post-onset. Dots indicate concordant directionality; crosses indicate discordance. Red, blue, and gray denote significance (q < 0.05) in both, one, or neither cohort, respectively.

**Supplementary Fig. 4:**
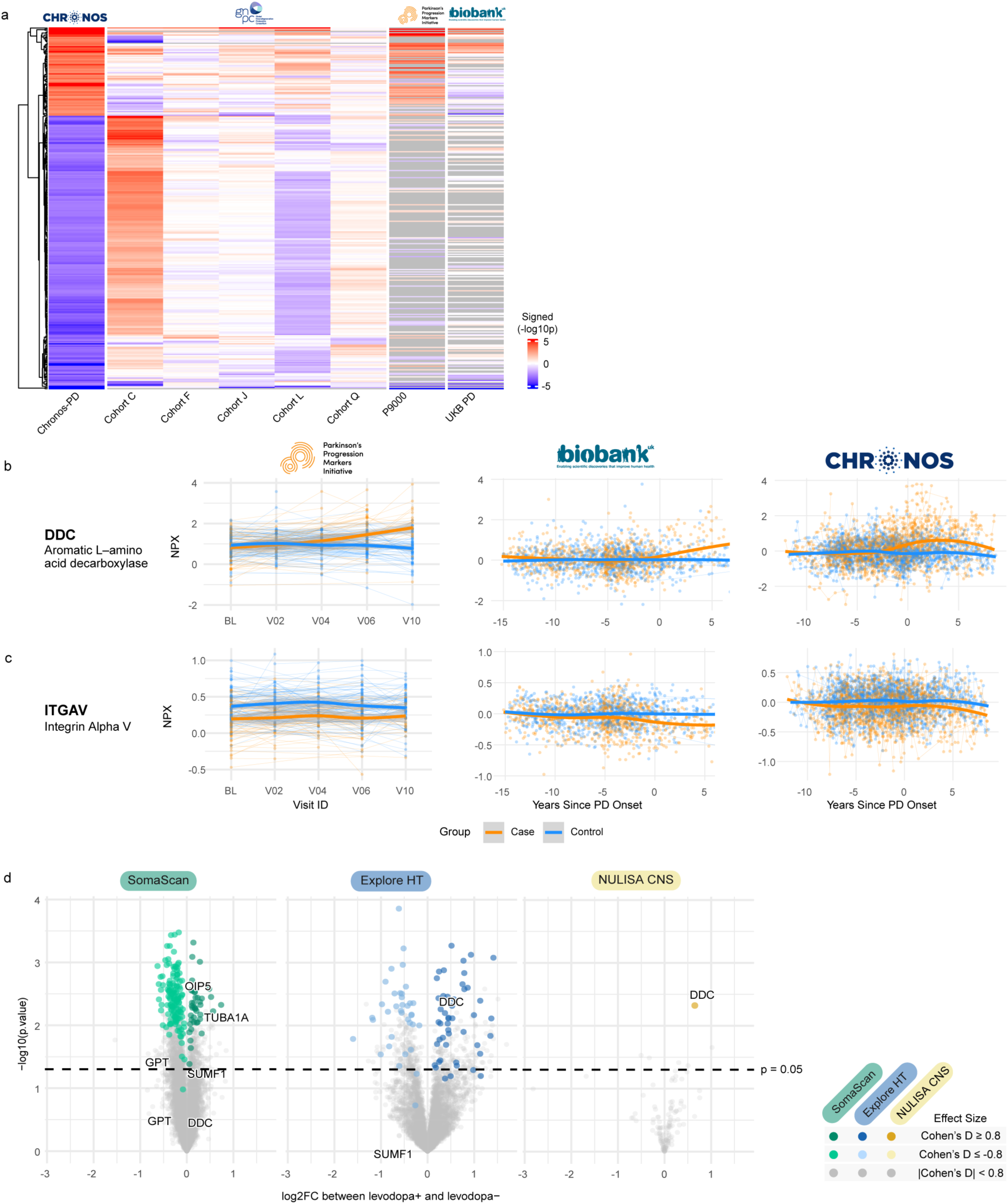
Extended validation of PD signals in external cohorts and proteomic effects of levodopa. **a**, Heatmap displaying the signed significance of proteins associated with PD across Chronos-PD, GNPC cohorts (C, F, J, L, Q), PPMI, and UK Biobank. Proteins not measured by a given platform are shown in gray. Red-orange represents increased proteins and blue-purple represents decreased proteins. **b**, Visualization of DDC levels measured by Olink in PPMI (78 PD and 129 controls, *n*_samples_ = 526), UK Biobank (923 PD and 923 controls, *n*_samples_ = 1,855), and Chronos-PD (348 PD and 348 controls, *n*_samples_ = 2,584), stratified by PD status. Longitudinal samples from the same individuals are connected. Dots indicate individual samples, thin lines connect samples from the same individuals, and thick lines were estimated mean trend over time in PD and control groups based on LOESS method (locally estimated scatterplot smoothing). **c**, Visualization of ITGAV levels measured by Olink in PPMI, UK Biobank, and Chronos-PD. Visualization and annotation are as in b. **d**, Volcano plots showing the effect of levodopa treatment across affinity proteomic platforms. Chronos-PD samples with levodopa prescription claims within one month prior to the sample collection date (levodopa+, *n*_case_ = 29, *n*_samples_ = 55) were compared with samples that had available medication data but no levodopa-containing prescriptions in the preceding year (levodopa-, *n*case = 25, *n*_samples_ = 58).

**Supplementary Fig. 5:**
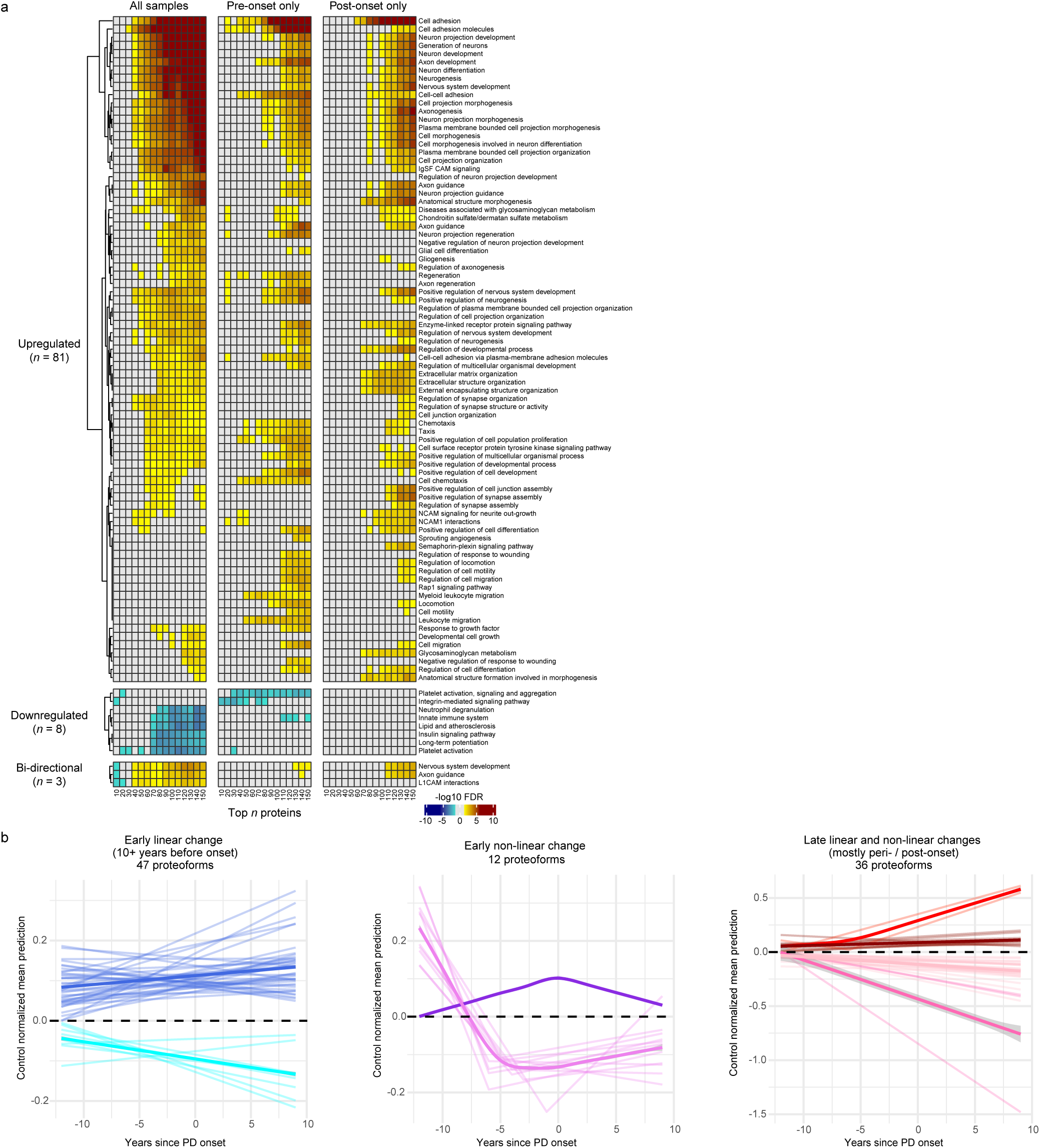
Expanded pathway and protein trajectories analysis. **a**, Expanded view of Fig. 3c, showing key enriched pathways among PD signals, from pre-onset, post-onset, and all samples, detected by SEPA. Brown represents upregulated pathways and blue represents downregulated pathways. **b**, Longitudinal trajectories of candidate biomarkers. Modeled levels of identified proteins based on linear and non-linear mixed-effects models, integrated with time to estimated PD onset via joint modeling. Protein levels are visualized as control-normalized estimates from 12 years pre- to 9 years post-onset, separated into three groups: early linear change, early non-linear change, or late linear and non-linear change. Individual protein trajectories are displayed, with thicker lines representing the average trajectories within each group, shown separately for proteins increasing or decreasing in PD.

**Supplementary Fig. 6:**
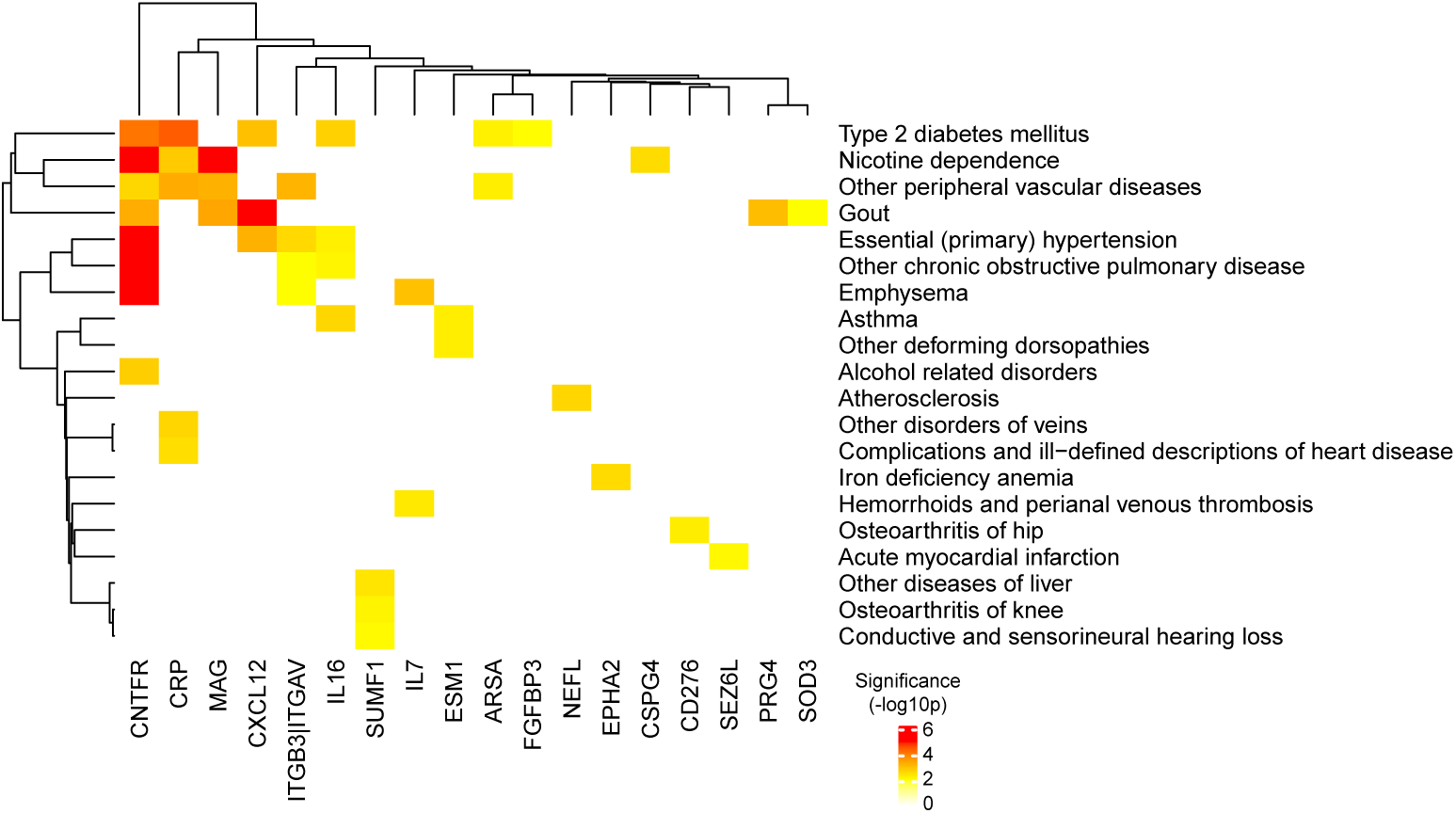
Expanded comorbidity analysis. Heatmap of significant associations between non-PD ICD-10 comorbidities and PD early/risk biomarkers (filtered to p > 0.01). Color corresponds to statistical significance for each association.

**Supplementary Fig. 7:**
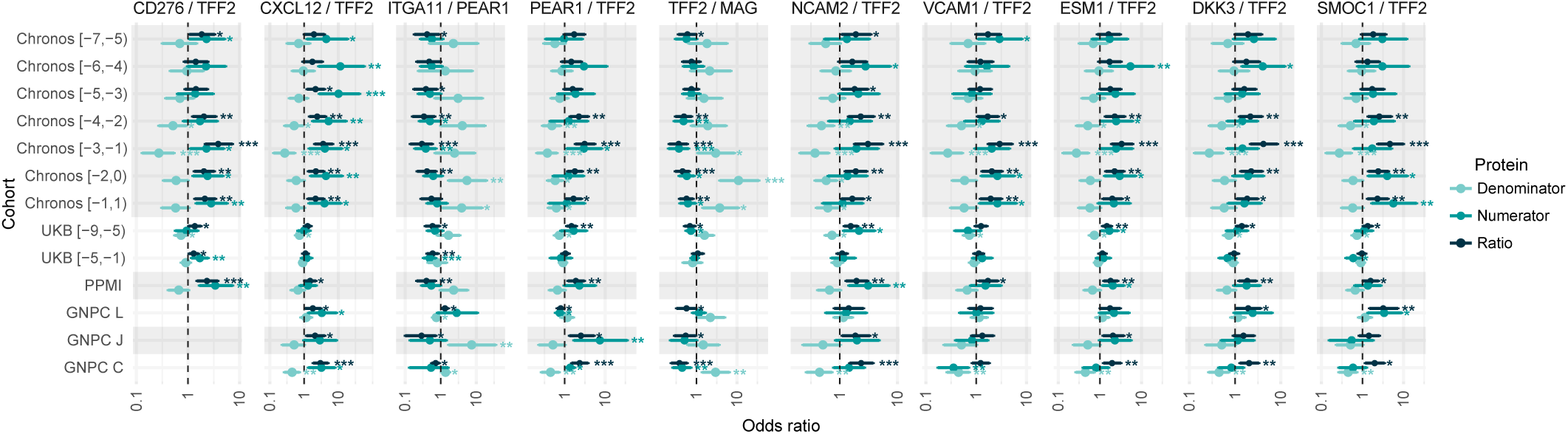
Comparison of protein ratios and their individual components. Forest plot of odds ratios for selected protein ratios (dark teal) and individual ratio members (light teal, teal) across cohorts. Points and error bars denote ORs and 95% CIs; vertical dashed lines indicate OR = 1. Significance levels: *p < 0.05, **p < 0.01, ***p < 0.001.

**Supplementary Fig. 8:**
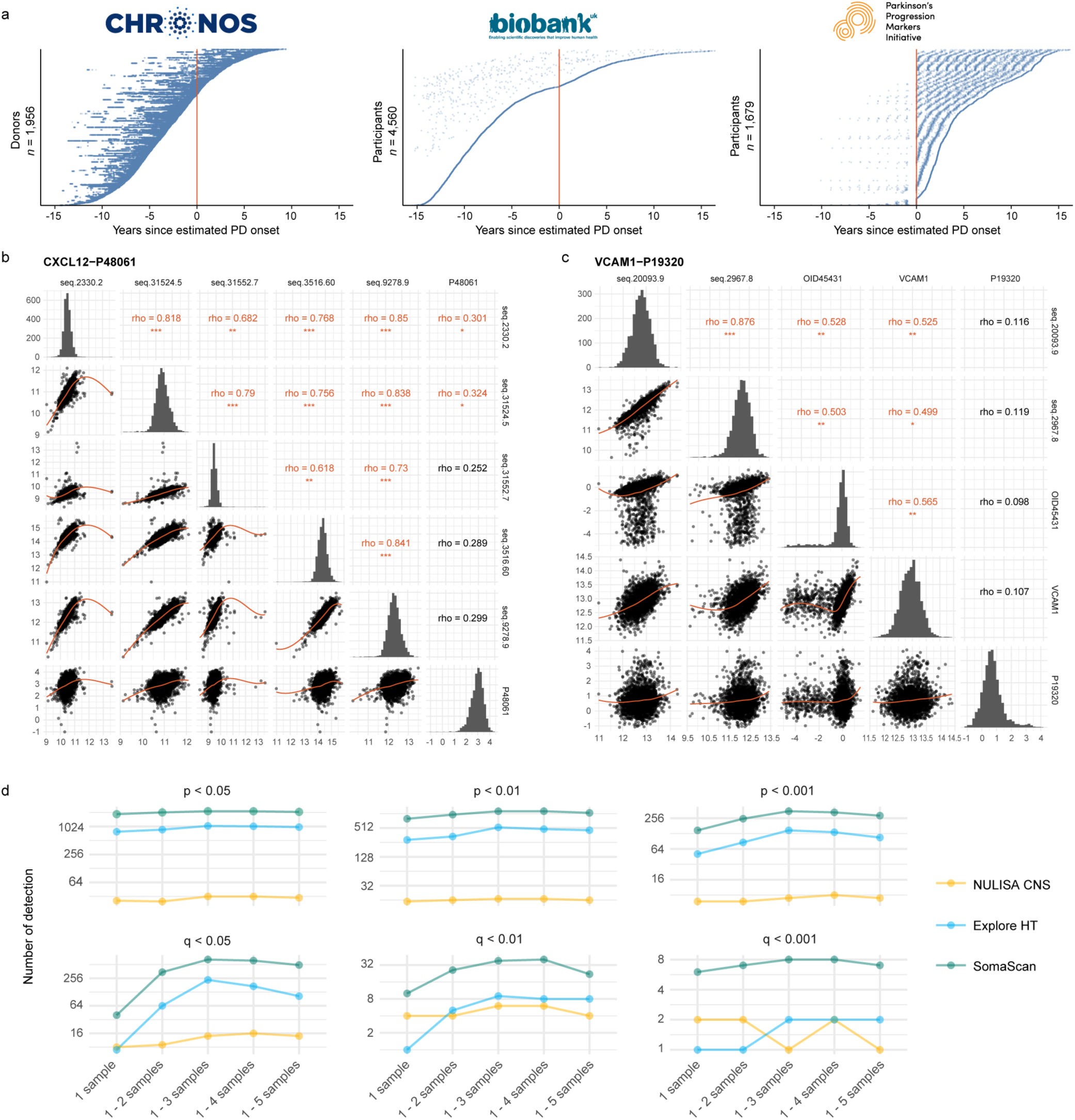
Biospecimen depth, cross-platform consistency, and longitudinal power. **a**, Distribution of plasma samples as a function of PD onset for Chronos-PD, UK Biobank, and PPMI cohorts. Plots are centered on the time of PD diagnosis or estimated PD onset, spanning from -15 to +15 years for comparative analysis. A total of 69 UK Biobank samples fall outside this temporal window and are not displayed. **b**, CXCL12 levels measured by 4 SOMAmers and Discovery MS. Rho is the spearman correlation: *0.3 < rho < 0.5, **0.5 < rho < 0.7, ***rho > 0.7. **c**, VCAM1 levels measured by 2 SOMAmers, Explore HT, NULISA CNS, and Discovery MS. Rho is the spearman correlation: *0.3 < rho < 0.5, **0.5 < rho < 0.7, ***rho > 0.7. **d**, Number of proteins significantly associated with PD per platform, comparing analyses using one sample per individual versus up to five longitudinal samples across different p-value and q-value thresholds given the same number of individuals.

**Supplementary Fig. 9:**
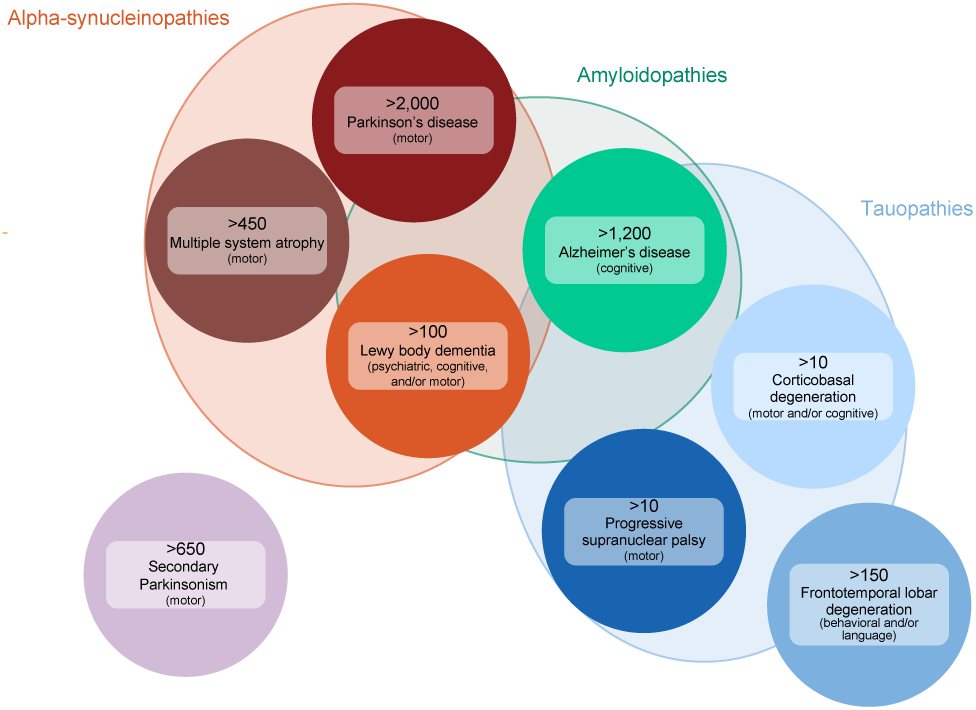
Number of individuals with PD and CNS diseases close to PD. Number of individuals with selected diseases overlapping with PD based on claims in the current Chronos registry.

